# Changes in symptomatology, re-infection and transmissibility associated with SARS-CoV-2 variant B.1.1.7: an ecological study

**DOI:** 10.1101/2021.01.28.21250680

**Authors:** Mark S. Graham, Carole H. Sudre, Anna May, Michela Antonelli, Benjamin Murray, Thomas Varsavsky, Kerstin Kläser, Liane S. Canas, Erika Molteni, Marc Modat, David A. Drew, Long H. Nguyen, Lorenzo Polidori, Somesh Selvachandran, Christina Hu, Joan Capdevila, The COVID-19 Genomics UK (COG-UK) consortium, Alexander Hammers, Andrew T. Chan, Jonathan Wolf, Tim D. Spector, Claire J. Steves, Sebastien Ourselin

## Abstract

**Background:** SARS-CoV-2 variant B.1.1.7 was first identified in December 2020 in England. It is not known if the new variant presents with variation in symptoms or disease course, if previously infected individuals may become reinfected with the new variant, or how the variant’s increased transmissibility affects measures to reduce its spread.

**Methods:** Using longitudinal symptom reports from 36,920 users of the COVID Symptom Study app testing positive for Covid-19 between 28 September and 27 December 2020, we performed an ecological study to examine the association between the regional proportion of B.1.1.7 and reported symptoms, disease course, rates of reinfection, and transmissibility.

**Findings:** We found no evidence for changes in reported symptoms or disease duration associated with B.1.1.7. We found a likely reinfection rate of 0.7% (95% CI 0.6-0.8), but no evidence that this was higher compared to older strains. We found an increase in R(t) by a factor of 1.35 (95% CI 1.02-1.69). Despite this, we found that R(t) fell below 1 during regional and national lockdowns, even in regions with high proportions of B.1.1.7.

**Interpretation:** The lack of change in symptoms indicates existing testing and surveillance infrastructure do not need to change specifically for the new variant, and the reinfection findings suggest that vaccines are likely to remain effective against the new variant.

**Funding:** Zoe Global Limited, Department of Health, Wellcome Trust, EPSRC, NIHR, MRC, Alzheimer’s Society.

**Research in context:** *Evidence before this study:* To identify existing evidence on SARS-CoV-2 variant B.1.1.7 we searched PubMed and Google Scholar for articles between 1 December 2020 and 1 February 2021 using the keywords Covid-19 AND B.1.1.7, finding 281 results. We did not find any studies that investigated B.1.1.7-associated changes in the symptoms experienced, their severity and duration, but found one study showing B.1.1.7 did not change the ratio of symptomatic to asymptomatic infections. We found six articles describing laboratory-based investigations of the responses of B.1.1.7 to vaccine-induced immunity to B.1.1.7, but no work investigating what this means for natural immunity and the likelihood of reinfection outside of the lab. We found five articles demonstrating the increased transmissibility of B.1.1.7.

*Added value of this study:* To our knowledge, this is the first study to explore changes in symptom type and duration, as well as community reinfection rates, associated with B.1.1.7. The work uses self-reported symptom logs from 36,920 users of the COVID Symptom Study app reporting positive test results between 28 September and 27 December 2020. We find that B.1.1.7 is not associated with changes in the symptoms experienced in Covid-19, nor their duration. Building on existing lab studies, our work suggests that natural immunity developed from previous infection provides similar levels of protection to B.1.1.7. We add to the emerging consensus that B.1.1.7 exhibits increased transmissibility.

*Implications of all the available evidence:* Our findings suggest that existing criteria for obtaining a Covid-19 test in the community need not change for the rise of B.1.1.7. The fact that immunity developed from infection by wild type variants protects against B.1.1.7 provides an indication that vaccines will remain effective against B.1.1.7. R(t) fell below 1 during the UK’s national lockdown, even in regions with high levels of B.1.1.7, but further investigation is required to establish the factors that enabled this, to facilitate countries seeking to control the spread of B.1.1.7.

## Introduction

In early December 2020, a phylogenetically distinct cluster of SARS-CoV-2 was genetically characterised in the South-East of England. The majority of cases had been detected in November with a small number detected as early as September.^1^ Genomic surveillance reveals that this new variant, termed B.1.1.7, has a number of mutations of immunologic significance and is growing rapidly in frequency and spread^2^. It is important that we understand how these mutations may affect the presentation and spread of Covid-19, so that we can formulate effective public health responses.^3^

Preliminary evidence from epidemiological studies suggests the new strain is more transmissible. Davies et. al. found the new strain is 56% (95% CI 50-74) more transmissible^4^ and Volz et. al. found the new strain increases the effective reproduction number R(t) by a factor of 1.4-1.8^5^. There is evidence to suggest B.1.1.7 increases risk of hospitalisation and death.^6^ However, there is much that is still unknown. From a public health perspective, it is crucial to understand if B.1.1.17 necessitates changes to existing measures for disease monitoring and containment. For instance, changes to symptomatology could require modifications to symptomatic testing programmes to ensure new cases are identified, and changes to disease duration could require changes in the amount of time infected individuals are required to isolate for. It is important for modelling and forecasting models to understand if B.1.1.7 alters the rate of reinfection. Early estimates of the new transmissibility of B.1.1.7 are uncertain and there is a need for additional estimates using independent data sources; furthermore it is important to understand how these findings will affect measures to control the pandemic’s spread using non-pharmaceutical interventions, such as lockdowns.

We make use of data from the COVID Symptom Study (CSS)^7^ to investigate the symptomatology, disease course, and transmissibility of the new variant. The longitudinal dataset provides symptom reports and test results from a population of over 4 million adults living in the UK using the mobile application. By combining these data with surveillance data from the Covid-19 UK Genetics Consortium (COG-UK)^8^ and a spike-gene target failure correlate in community testing data, we performed associative, ecological studies to assess the symptoms, disease course, rates of reinfection, and transmissibility of the new variant.

## Methods

### Symptom study data

Longitudinal data were prospectively collected using the CSS app, developed by Zoe Global with input from King’s College London (London, UK), the Massachusetts General Hospital (Boston, MA, USA), and Lund and Uppsala Universities (Sweden). The app guides users through a set of enrolment questions, establishing baseline demographic and health information. Users are asked to record each day whether they feel physically normal, and if not, to log any symptoms. After a user reports any symptoms, they are asked “Where are you right now?”, with the options “At home”, “At hospital with suspected Covid-19 Symptoms”, or “Back from hospital”. Users are also asked to maintain a record of any Covid-19 tests, their date, type, and result in the app. Users are able to record the same data on behalf of others, such as family members, to increase data coverage amongst those unlikely to use mobile applications, such as the elderly. More details about the app can be found in a study by Drew and colleagues.^7^ We included users living in the UK who had logged in the app at least once in the period between 28 September to 27 December 2020.

### Genomic data

We used data released on 13 January 2021 from COG-UK to extract time-series of the percentage of daily cases that came from the B.1.1.7 lineage in Scotland, Wales, and each of the seven National Health Service (NHS) regions in England. Northern Ireland was excluded due to the low number of samples in the COG-UK dataset. These data are produced by sequencing a sample of polymerase chain reaction (PCR) tests carried out in the community. Due to the delay of approximately two weeks^2^ between PCR and genomic sequencing, we only used data from samples taken up to 31 December to avoid censoring effects.

Additionally, we used data from Public Health England (PHE) on the probable new variant captured in community cases in England using spike gene target failure (SGTF). It has been observed that one of the spike gene mutations in B.1.1.7 causes an SGTF in the test used in three of England’s large laboratories used for analysis of community cases.^1^ This failure results in a marker that is sensitive to B.1.1.7, but not necessarily specific, as other circulating variants also contain the mutation leading to an SGTF. Comparison to genomic data finds that from 30 November 2020 onwards more than 96% of cases with the SGTF were from lineage B.1.1.7.^9^ The proportion of SGTF cases is made available in England for each of the 316 “Lower Tier” Local Authorities. We grouped these data to each NHS region using a population-weighted average to enable integration with other data sources.

### Disease symptoms and course

In order to assess whether the symptomatology of infection from B.1.1.7 differed from previous variants, we investigated the change in symptom reporting from 28 September to 27 December 2020, covering 13 complete weeks over the period when the proportion of B.1.1.7 grew most notably in London, South East and East of England. For each week in every region considered, we calculated the proportion of users reporting each symptom. Users were included in a week if they had reported a positive swab test (PCR or lateral flow) in the period 14 days before or after that week. For each region and symptom we performed a linear regression, examining the association between the proportion of B.1.1.7 in that region (independent variable) and the proportion of users reporting the symptom (dependent variable) over the 13 weeks considered. We adjusted for the age and sex of users, as well as two seasonal environmental confounders: regional temperature and humidity. Seasonal confounders were calculated each day the temperature and relative humidity at two meters above the surface, averaged across each region considered.^10^

We also examined the association between proportion of B.1.1.7 and disease burden, measured here as the total number of different symptoms reported over a period of two weeks before and two weeks after the test, and the relation with asymptomatic infection, defined as users reporting a positive test result but no symptoms in the two weeks before or after the test. We also investigated the rate of self-reported hospital visits, including both users who reported being in hospital with suspected Covid-19 symptoms, and being back from hospital. We also investigated the proportion of individuals reporting long symptom duration using a previously published definition of continuous symptoms reported for at least 28 days.^11^ To avoid censoring effects, both hospitalisation and long duration analyses included symptom reports extended up to 18 January, and the long duration analysis only considered reports of positive tests up to 21 December. All analyses adjusted for sex, age, temperature and humidity. We controlled for the false discovery rate to account for multiple comparisons.

### Reinfection

We defined possible reinfection as the presence of two reported positive tests separated by more than 90 days with a period of reporting no symptoms for more than seven days before the second positive test. We calculated the proportion of possible reinfections among individuals reporting their first positive test before 1 October 2020. To assess whether the risk of reinfection was stronger in the presence of the new variant, in every region we performed ecological studies, examining the Spearman correlation between the proportion of B.1.1.7 cases and the number of reinfections over time, and between the proportion of positive tests reported through the app and the number of reinfections. We compared these two correlations in each region using the Mann-Whitney U test.

### Transmissibility

Daily incidence for Scotland, Wales, and each of the seven NHS regions in England were produced from the period 1 October 2020 to 27 December 2020 using data from the CSS app and previously described methodology.^12^ Using the COG-UK data to estimate the proportion of B.1.1.7 in circulation in each region per day, these incidence estimates were decomposed into two incidence time-series per region, one for the old variants and and one for B.1.1.7, with the constraint that the two time-series should sum to match the total incidence. R(t) was estimated separately for the old and new variants using previously described methodology; briefly, we used the relationship I_t+1_=I_t_□exp(μ□(R(t)□–□1), where 1/μ is the serial interval and I_t_ the incidence on day t. We modelled the system as a Poisson process and assumed that the serial interval was drawn from a gamma distribution with α=6·0 and β=1·5, and used Markov Chain Monte-Carlo to estimate R(t). We compared both multiplicative and additive differences of the new and old R(t) values for days when the proportion of B.1.1.7 in a region was greater than 3%. While data is not available for the proportion of B.1.1.7 in January, we also computed total incidence and R(t) from 1 October 2020 to 16 January 2021 to see how they changed during national lockdown in England.

## Results

### Symptom study data

Table 1 shows the demographic data for the cohort studied. From 24 March to 27 December 2020, 4,327,245 participants from the UK signed up to use the app. We excluded users living in Northern Ireland due to the low number of sign-ups (38,976), 383,352 users lacking information on sex, and 2,175,979 who had not logged in the app during the period 28 September to 27 December 2020, leaving a total of 1,767,914 users. Between them, these users recorded 65,606,869 logs in the app between 28 September and 27 December. In this period, 497,989 users reported a swab test. 55,192 of these reported a positive test, and we investigated the symptom reports of 36,920 of those whose region was known and who reported as healthy on app sign-up.

**Table 1.** Characteristics of GB app users active in the period 28 September − 27 December 2020 * Reports logged between 28 September - 27 December. For some analyses we took further reports from an extended time period 14 September 2020 - 18 January 2021 **May be more than one test per individual as the overall number contains failed tests and unknown results

|  |  | Overall |  | Tested |  | Tested positive** |  | Signed up healthy with reporting around positive test |  |
| --- | --- | --- | --- | --- | --- | --- | --- | --- | --- |
|  |  | N | % | N | % | N | % | N | % |
| **Users** |  | 1,767,914 | --- | 497,989 | --- | 55,192 | --- | 40,463 | --- |
| **Daily reports\*** |  | 65,613,697 | --- | 19,154,601 | --- | 1,514,244 | --- |  | --- |
| **Age in years mean (std)** |  | 48.4 (19.3) | --- | 46.06 (17.8) | --- | 42.1 (16.8) | --- | 42.9 (17.0) | --- |
|  | ≤18 | 163,112 | 9.2 | 40,717 | 8.2 | 5,468 | 9.9 | 3,874 | 9.6 |
|  | 19 - 64 | 1,234,259 | 69.8 | 381,900 | 76.7 | 45,149 | 81.8 | 32,878 | 81.2 |
|  | ≥ 65 | 370,543 | 20.9 | 72,741 | 14.6 | 4,367 | 7.9 | 3,600 | 8.9 |
|  | Invalid | 5,576 | 0.3 | 2,631 | 0.5 | 208 | 0.3 | 111 | 0.3 |
| **Sex** | Female | 1,046,074 | 59.2 | 315,875 | 63.4 | 34,516 | 62.5 | 24,844 | 61.4 |
|  | Male | 720,562 | 40.8 | 181,110 | 36.4 | 20,546 | 37.2 | 15,545 | 38.4 |
|  | Intersex | 79 | <0.1 | 21 | <0.1 | 3 | <0.1 | 3 | <0.1 |
|  | Prefer not to say | 1,199 | 0.1 | 983 | 0.2 | 127 | 0.2 | 71 | 0.2 |
| **Region** | South East | 342,881 | 19.4 | 97,143 | 19.5 | 8,762 | 16.0 | 6,555 | 16.2 |
|  | East of England | 196,063 | 11.1 | 57,680 | 11.6 | 5,373 | 9.8 | 4,037 | 10.0 |
|  | London | 227,004 | 12.8 | 81,940 | 16.5 | 9,733 | 17.8 | 7,384 | 18.2 |
|  | Midlands | 198,350 | 11.2 | 57,582 | 11.6 | 6,695 | 12.2 | 4,756 | 11.8 |
|  | North East and Yorkshire | 156,999 | 8.9 | 42,986 | 9.1 | 5,292 | 9.7 | 3,744 | 9.3 |
|  | North West | 123,201 | 7.0 | 45,156 | 9.1 | 6,180 | 11.3 | 4,399 | 10.9 |
|  | South West | 186,372 | 10.5 | 46,780 | 9.4 | 3,685 | 6.7 | 2,637 | 6.5 |
|  | Scotland | 87,263 | 4.9 | 13,793 | 2.8 | 1,589 | 2.9 | 1,049 | 2.6 |
|  | Wales | 82,886 | 4.7 | 16,471 | 3.3 | 3,092 | 5.6 | 2,359 | 5.8 |
|  | Not known | 165,164 | 9.3 | 38,458 | 7.5 | 4,638 | 8.0 | 3,543 | 8.8 |

### Genomic data

In the period between 27 September 2020 and 31 December 2020, 98,170 sequences were made available by COG-UK, corresponding to 4.4% of the 2,207,476 cases recorded in this period.^13^ 16,224 sequences (16.5%) were variant B.1.1.7. Considering the mean of the rolling average across December, the three regions with the largest proportion of B.1.1.7 are the South East, London, and East of England. The three regions with the lowest proportion are Wales, the North East and Yorkshire, and the North West. SGTF data was made available in England on a weekly basis from 10 November 2020 to 29 December 2020. Of the 700,590 cases reported in this period, 295,404 (42.2%) caused an SGTF. Examining the COG-UK data from England in the same time period, we find 34.6% cases are B.1.1.7. The difference is in part attributable to the SGTF being a nonspecific marker of B.1.1.7: in the week from 9-15 November 81% of cases with an SGTF were B.1.1.7, while from 30 November at least 96% of cases with the SGTF were from B.1.1.7. Figure 1 shows how the proportion of the new variant changed over time in regions of the UK using COG-UK and the SGTF data.

**Figure 1.**
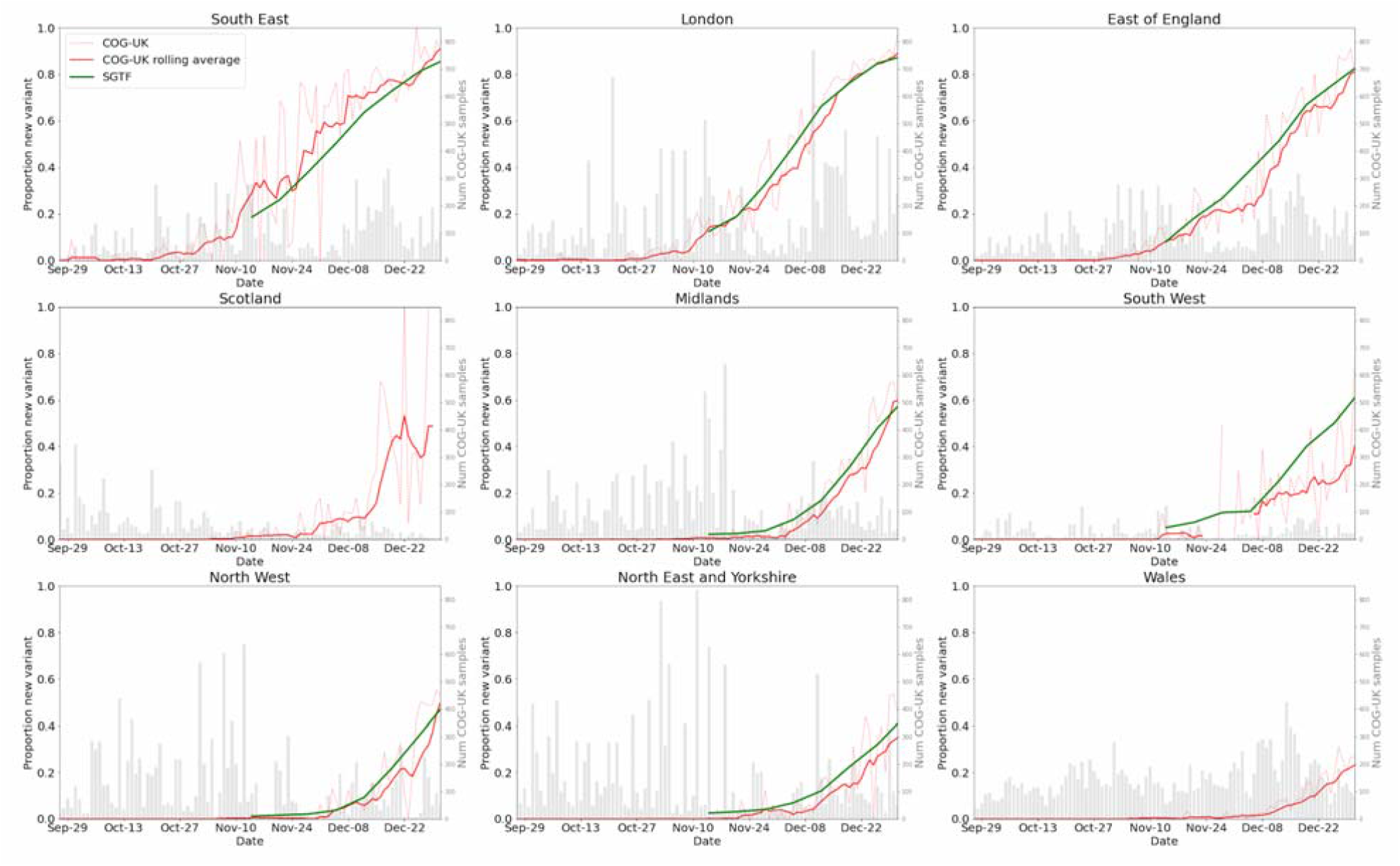
Presence of B.1.1.7 in each of the 7 NHS regions in England, and Scotland and Wales, as measured using genomic surveillance data (COG-UK) and SGTF data. SGTF data are not available for Scotland or Wales.

### Disease symptoms and course

Figure 2 illustrates the variation of symptom occurrence over time considering a one-week window smoothed over 3 time points as a function of time, and Supplementary Figure 1 shows how these symptoms vary as a function of the proportion of B.1.1.7. Qualitatively, these results show no change in the proportion of users reporting each symptom with the new variant. Linear regression (both unadjusted and adjusted for age, sex, temperature and humidity) did not find evidence of association between the proportion of B.1.1.7 and symptoms reported after controlling the false discovery rate. Both adjusted and unadjusted results are shown in Supplementary Figure 4.

**Figure 2.**
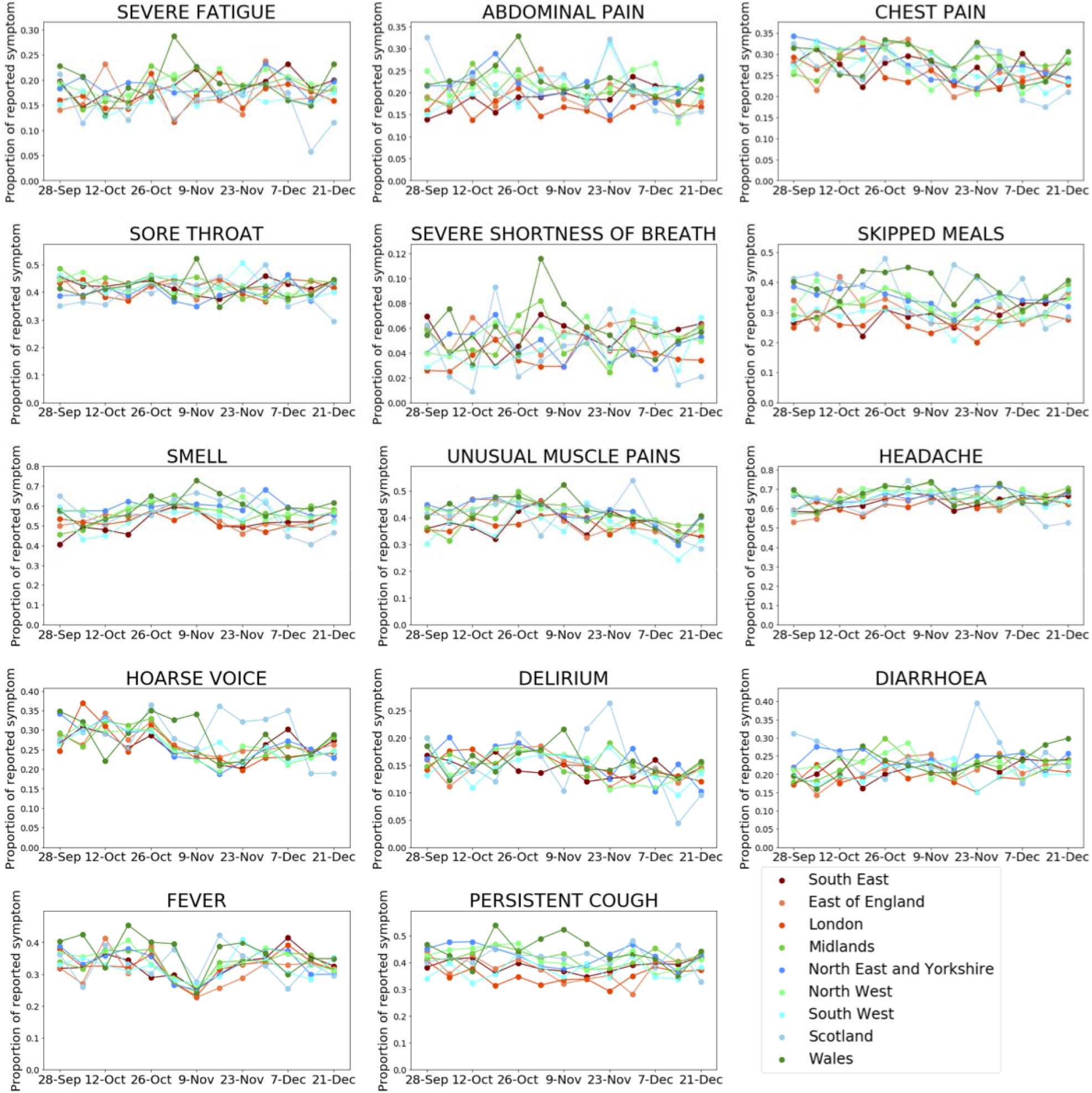
Regional plots of the frequency of reporting of symptoms over time for each reported symptom. Drop in fever reporting in early November was caused by a change in the question wording; this wording was subsequently reverted a week later.

Figure 3 shows the variation of total number of symptoms reported, the total number of asymptomatic infections, self-reported hospital visits, and instances of long symptom duration over time, and Supplementary Figure 2 shows how these plots vary with proportion of B.1.1.7. Visually, we see no change in any of these outcomes with an increasing proportion of B.1.1.7. When correcting for mean age, sex, ambient temperature and humidity there was no evidence of an association between B.1.1.7 and either the number of symptoms reported over a 4-week window, the number of hospitalisations, long symptom duration, or proportion of asymptomatic cases, see Supplementary Table 1.

**Figure 3.**
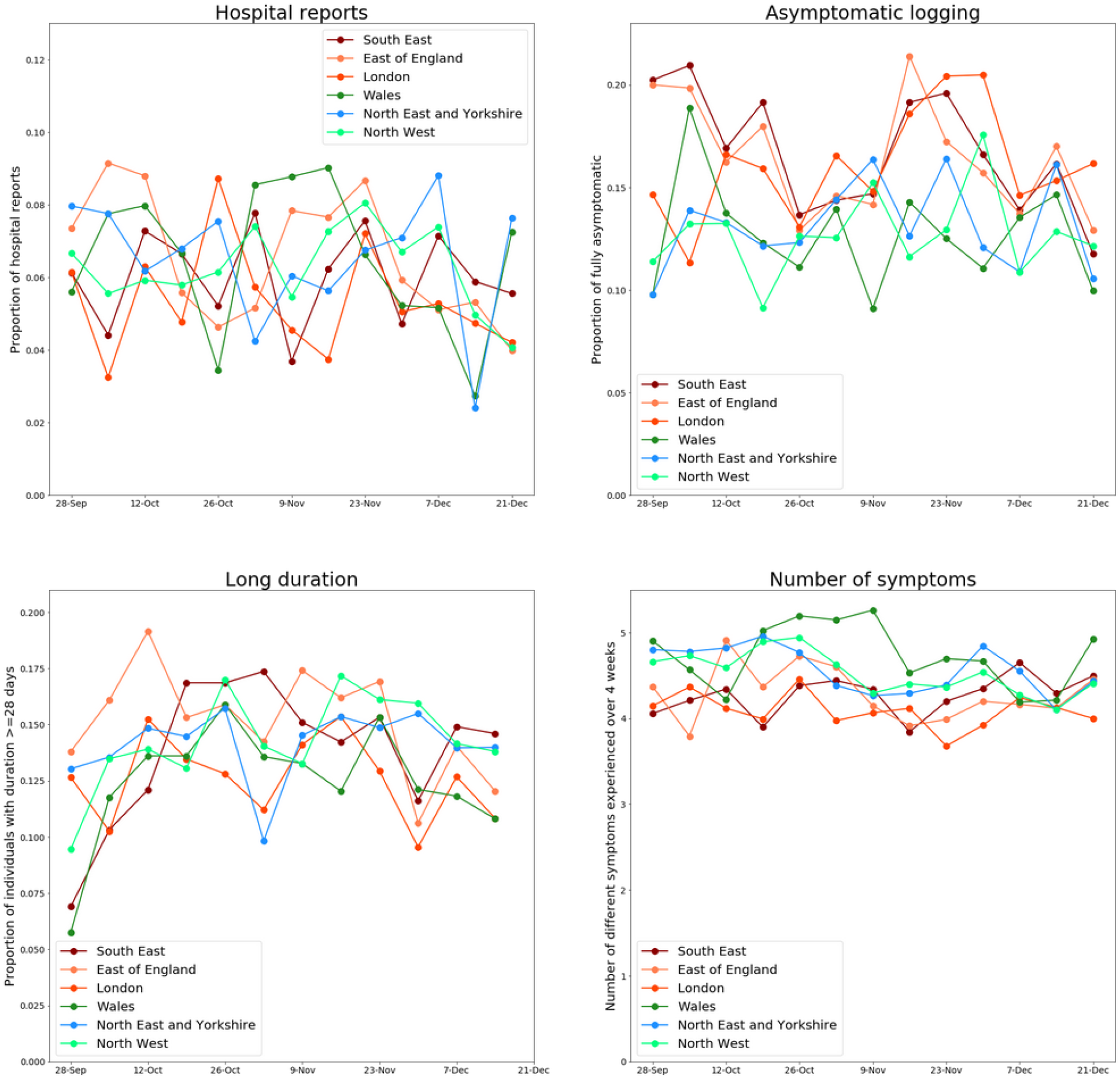
Regional plot of hospitalisation reports, proportion of asymptomatic positives, instances of long symptom duration and the total number of different symptoms reported. For the study of long symptom duration, tests are only considered up to 21 December, symptom reports up to 18 January 2021 to limit right censoring effects. Only symptomatic individuals for which duration can be ascertained are included.

### Reinfection

Overall, we identified 304 individuals reporting two positive tests with more than 90 days between the two. Among these individuals, symptom reporting allowed us to identify 249 for which there is a period of at least 7 symptom-free days in between positive tests among the 36,509 individuals having reported a positive swab test before 1 October 2020 (0.7%, 95% CI 0.6-0.8). Among those 249, daily reports were available in the periods around both of the positive tests for 173. There was no difference in reinfection reporting rates across the different NHS regions (p=0.1). Figure 4 shows the evolution in the number of possible reinfections along with reported positive cases (green line) and proportion of B.1.1.7 (red line). For all regions (except Scotland), reinfection occurrences were more positively correlated with the overall regional rise in cases rather than the regional rise in the new variant percentage (Number of cases:reinfection, Spearman rho 0.55 to 0.69 [p<0.05] for South East, London and East of England; % new variant:reinfection, Spearman rho 0.37 to 0.55 in the same regions). Supplementary Table 2 shows the bootstrapped median values of correlation compared across the different regions and the outcome of a Mann-Whitney U test across the bootstrapped distributions.

**Figure 4.**
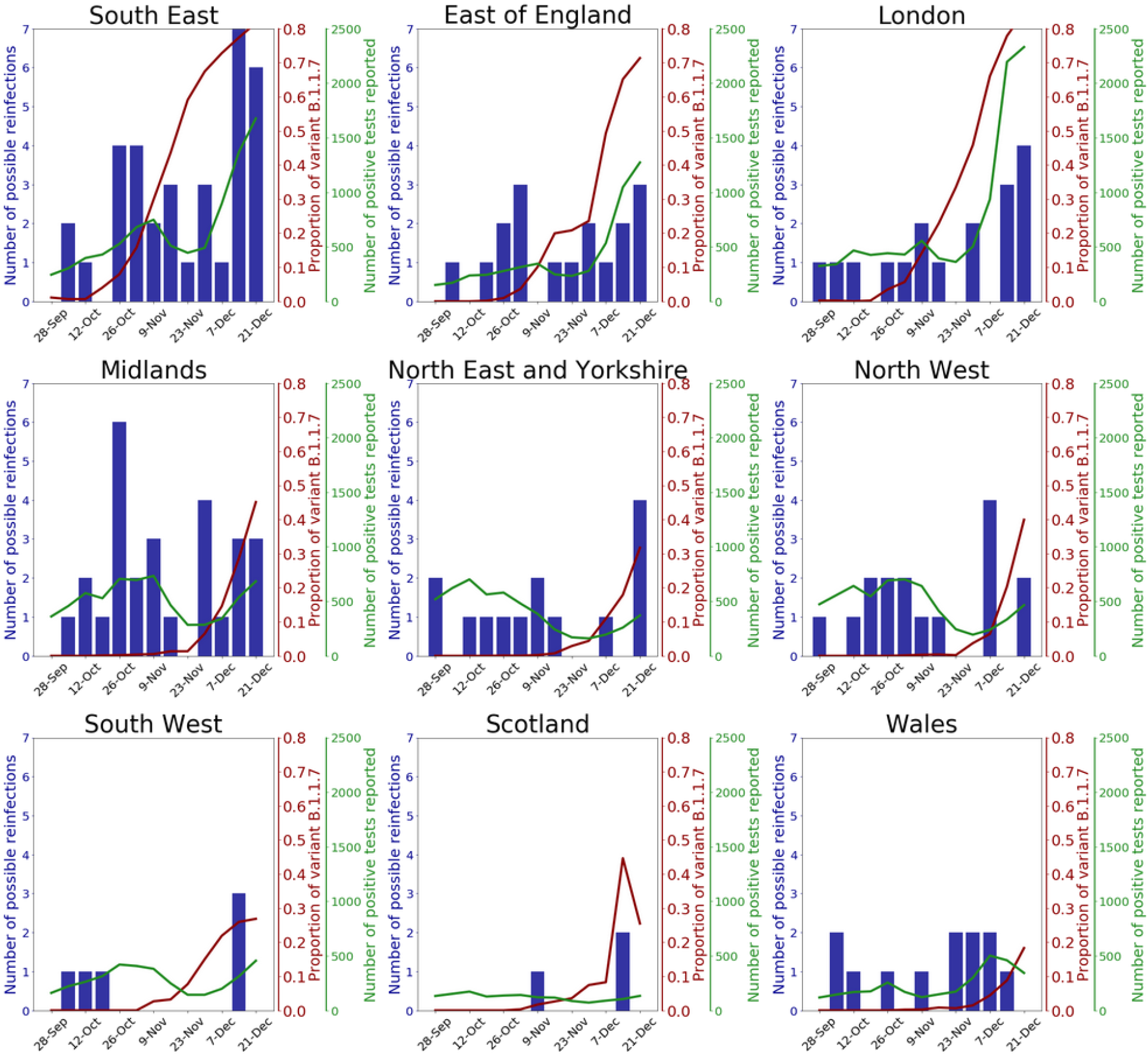
Number of reinfection reports by region according to week of the second infection, along with the total number of positive tests reported through the app (green line) and the proportion of B.1.1.7 in circulation (red line).

### Transmissibility

Figure 5 shows incidence and R(t) for the old and new variants in the three regions in England with the highest proportions of the new variant. Results consistently show the R(t) of B.1.1.7 to be greater than that of other variants. The mean (95% CI) of the additive increase in R for B.1.1.7 was 0.34 (0.02-0.66), and the multiplicative increase was 1.35 (1.02-1.69). England exited its second national lockdown on 2 December, leading to a change in behaviour and R(t). When considering only the period after the second lockdown ended, we find 0.28 (0.01-0.61) for the additive and 1.28 (1.02-1.61) for the multiplicative increases. Supplementary Figure 3 shows the same using the SGTF data, with analysis limited to the period after 1 December when at least 95% of all SGTF cases were B.1.1.7. These data are provided weekly, and linear interpolation was used to obtain daily estimates, leading to smoother estimates for variant-specific incidence and R(t). Using these values, we find R(t) of B.1.1.7 has an additive increase of 0.26 (0.15-0.37) and a multiplicative increase of 1.25 (1.17-1.34).

**Figure 5.**
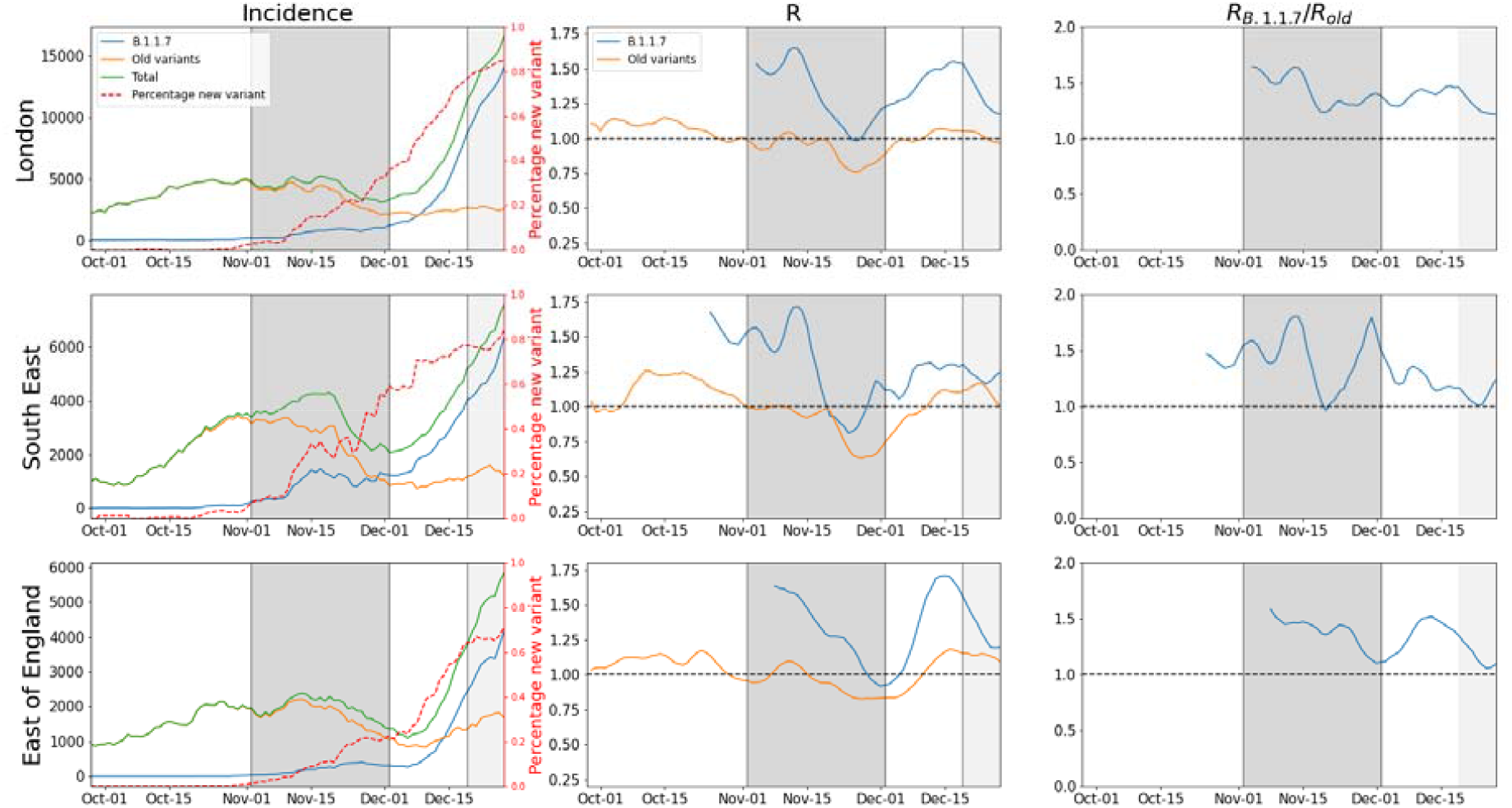
Incidence and R(t) for the old and new variants, along with the ratio between these R values, for the three regions in England with the largest proportion of B.1.1.7. Dark grey regions indicate national lockdowns, light grey the period where London and much of the South East and East of England were placed in Tier 4 restrictions.

On 19 December 2020 London and much of the South East and East of England were placed in ‘Tier 4’ restrictions, enforcing stricter rules for social distancing and decreased human-to-human contact that stopped short of nationwide measures. On 5 January 2021 the whole of England was placed in national lockdown. Figure 6 shows overall incidence and R(t) for the longer period from 1 October 2020 to 16 January 2021 in the three regions with the largest proportion of B.1.1.7. The proportion of B.1.1.7 in these regions in January is at least 80%, assuming the proportion has not decreased from the end of December. R(t) fell to ∼0.8 in all three of these regions during national lockdown. An extended plot including Scotland, Wales, and all regions in England, is shown in Supplementary Figure 5.

**Figure 6.**
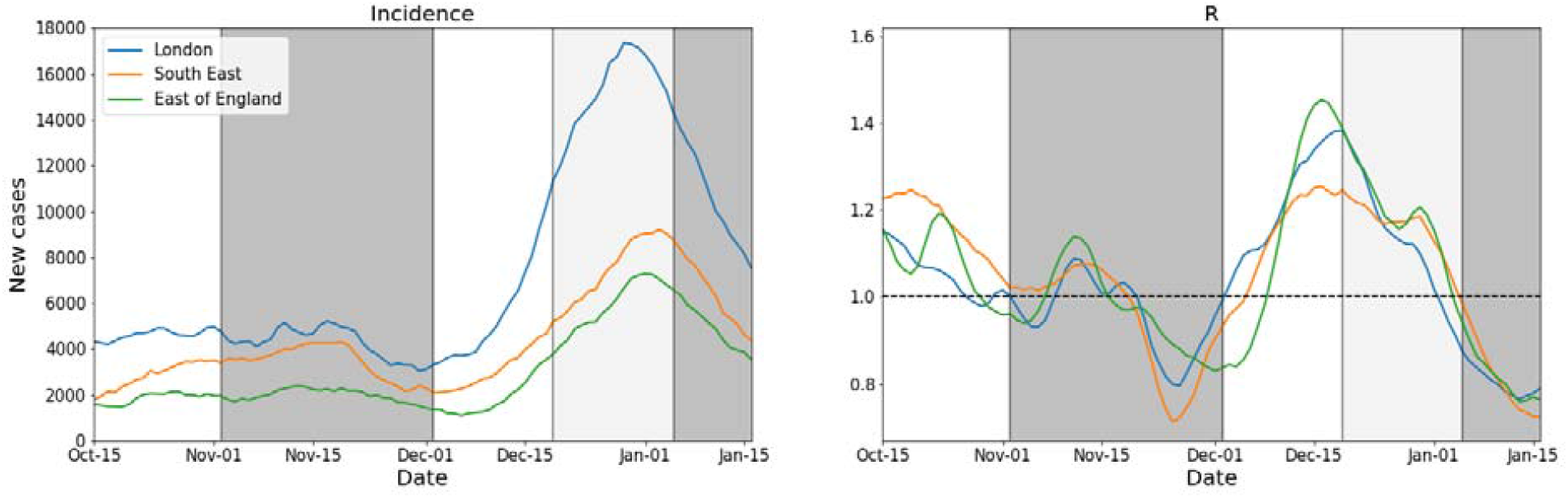
Total incidence and R(t) for the three regions with the highest proportion of B.1.1.7 in December, extended to capture the third national lockdown beginning 5 January 2021. Dark grey regions indicate national lockdowns, light grey indicate the period where London and much of the South East and East of England were placed in Tier 4 restrictions.

## Discussion

Using data collected through community reporting of symptoms and tests via the COVID Symptom Study app, we performed an ecological study to investigate whether the appearance of the variant B.1.1.7, first detected in a sample from England in September 2020, was associated with differences in symptoms experienced, disease duration, hospitalisation, asymptomatic infection, risks of reinfection, and transmissibility for users reporting a positive test result between 28 September and 27 December 2020.

We did not find associations between the proportion of B.1.1.7 in circulation and the type of symptoms experienced by our app users. We also did not find evidence for any change in the total number of symptoms experienced by individuals associated with B.1.1.7, nor the proportion of individuals experiencing long disease duration, defined as recording symptoms for more than 28 days without a break of more than seven days. We were also able to assess rates of asymptomatic disease and hospitalisation associated with B.1.1.7. We found the proportion of users with asymptomatic disease did not significantly change as B.1.1.7 increased in prevalence, in agreement with other work on the subject.^14^ We did not find any changes in hospitalisation, but other work has found evidence that B.1.1.7 increases hospitalisation rates.^6^ Limitations to the assessments of asymptomatic rates and hospitalisation should be noted: the majority of our users only get tested when they have symptoms and so we have relatively few asymptomatic infections recorded, and the self-reported nature of our hospitalisation data means we likely miss cases of more severe hospitalisation, when the individual is unlikely to self-report. There is also evidence that infection with B.1.1.7 carries increased risk of mortality;^6^ our data does not allow for us to assess this.

A recent study by the UK Office for National Statistics Community Infection Survey (CIS) reported that individuals infected with B.1.1.7 were more likely to report a cough, sore throat, fatigue, myalgia and fever in the seven days preceding the test, and less likely to report a loss of taste or smell.^15^ It is not clear if this report adjusted for age, sex, and environmental factors, though our analysis did not find this adjustment affected our findings (Supplementary Figure 4). The discrepancy may be explained by sampling at different points in the disease course. Our users predominantly seek testing at symptom onset, whilst the CIS design means the test may be administered at any point during the disease. Recent evidence has shown that B.1.1.7 causes longer infections^16^ which means the CIS symptom reports from users infected with B.1.1.7 may be sampled from later in the disease course than non-B.1.1.7 cases, causing apparent differences.The periods considered also differed: we considered symptoms reported both two weeks before and after the positive test result, compared to the one week before the positive test considered by the CIS. Further opportunity to study symptoms with B.1.1.7 in different contexts are required to be definitive.

We observed, based on 249 potential cases, a very low rate of possible reinfection of 0.7% (95% CI 0.6-0.8). This rate is consistent with another study of 6614 healthcare workers that had previously tested positive for Covid-19, finding 44 possible reinfections (0.66%).^17^ Our reinfection rate did not vary consistently across regions or time, which would be consistent with the hypothesis that reinfection is no more likely in the context of B.1.1.7. This may mean that if adequate immunity is built over the first infection it may be sufficient to protect against reinfection in the presence of B.1.1.7. Ultimately this is a positive sign that the immunity built through vaccination against the old variants could also be effective against B.1.1.7. This is in line with initial, laboratory-based studies regarding the efficacy of vaccines designed for early strains against this newer variant.^18–20^

We found an increase in the reproduction number R(t) in association with the B.1.1.7 variant: we found a multiplicative increase in R(t) of ∼1.35 (95% 1.02-1.69), compatible with estimates from Volz et al, of 1.4-1.8, Davies et al. who estimated a transmissibility increase of 1.56 (95% CI 1.50-1.74), and Walker et al who found an increase in growth rate that corresponds to a transmissibility increase of 1.33 (95% CI 1.21 - 1.53) if we assume a generation time of 4.7 days.^4,5,14,21^ These increases in transmissivity have worrying implications for the ability of lockdown measures to control B.1.1.7, given R(t) was estimated to be 0.7-0.9 during the first national lockdown in England.^22^ Despite this, we found R(t) to be ∼ 0.8 in the three regions in England with at least 80% of B.1.1.7. during the national lockdown beginning on 5 January 2021. There are several potential explanations for this. It could be that adherence to this lockdown is greater than previous lockdowns, helping to reduce R(t). It may also indicate the true increase in transmissivity is at the lower end of the available estimates, or that the increase in transmissivity estimated outside of lockdown cannot be extrapolated to lockdown, perhaps due to B.1.1.7 responding differently to lockdown measures than the old variants. Another possible explanation is that there is now sufficient community immunity to reduce R(t) further than seen during previous lockdowns. One serology study estimates that in the period 21 December 2020 - 18 January 2021, 15.3% (95% CI 14.7% to 15.9%) individuals in England would have tested positive for Covid-19 antibodies.^23^ Many countries have now detected cases of B.1.1.7 and work to better understand the factors that helped suppress it in the UK will help other countries to formulate their public health responses.^24^

Our study has several strengths. The large, longitudinal nature of the CSS data, with good coverage of the UK population, provides a unique opportunity to study potential changes in symptomatology and disease duration. The ability to match tests and symptom reports over long periods further allows us to measure possible reinfection rates. Our data also offers the ability to provide a valuable complementary measure to existing measurements of the increased transmissibility of B.1.1.7: we were able to use real-time, representative incidence estimates to measure R(t), whilst other studies have relied on deaths and hospitalisations, which are lagged, or community case numbers which do not reflect true infection numbers.

We acknowledge several limitations to this work. As we lack information on the disease strain of individual positive infections reported through the app, we performed an ecological study, assessing the association between the proportion of B.1.1.7 and population-level measures. This design does not allow for causal interpretation of the effect of B.1.1.7 on the measures we investigate. Our work assumes that all non-B.1.1.7 variants in circulation at the time of study give rise to the same symptomatology and immune response, and have the same transmissibility. Genomic surveillance has detected a very low number of non-B.1.1.7 variants of concern in circulation^25^, supporting the validity of this assumption, but it cannot be ruled out that other variants with different characteristics are circulating undetected. Data obtained from participatory, digital platforms have well-documented^26^ biases in demographics. Whilst we were able to correct for some of these factors in our analysis, such as age and sex, there are others that are more difficult to characterise and correct for. For example, respondents signing up to a participatory platform such as the CSS app are likely to be more interested in health and COVID-19 than the wider population, and may exhibit different behaviour. Participatory studies may also suffer from ascertainment or collider bias.^27^ Self report also carries the risk of data input errors, although the design of the app seeks to minimise this; for example, each time a user submits a log in the app they are shown the full history of their test results and are given the option to amend incorrect entries. Previous publications from our group have found that population-level estimates of disease prevalence from our app triangulate well with those obtained from studies designed to be representative of the population.^12^. We make the assumption that testing positive for SARS-CoV-2 after an interval of 90 days with at least a seven day period with an absence of symptoms is consistent with reinfection. Repeated positive testing has been reported shortly after hospital discharge^28^ and showed that PCR positivity could be detected up to 28 days post symptom resolution. While the chosen cut-off of 90 days between two positive tests is unlikely to be due to prolonged PCR positivity, this cannot be ruled out, but would only affect a small number of cases. Viral sequencing of the two infections would ideally be required to confirm reinfection. Despite correcting for changes in temperature and humidity, a possible limitation in the study is that comparisons in symptoms are made over time, and seasonal effects (e.g. on symptoms) may not have been fully taken into account.^29^

## Conclusions

We examined the effect of SARS-CoV-2 variant B.1.1.7 on the symptoms, disease course, rates of reinfection, and transmissibility in the UK. We found no change in symptoms or their duration. We found a low rate of reinfection (0.7%) and no evidence of increased rates associated with B.1.1.7. We found an increase in R(t) of ∼ 1.38 (95% CI 1.06-1.71), but evidence that R(t) fell below 1 during lockdown even in regions with very high (>80%) proportions of B.1.1.7.

## Data Availability

Data collected in the COVID Symptom Study smartphone application are being shared with other health researchers through the UK National Health Service-funded Health Data Research UK (HDRUK) and Secure Anonymised Information Linkage consortium, housed in the UK Secure Research Platform (Swansea, UK). Anonymised data are available to be shared with researchers according to their protocols in the public interest (https://web.www.healthdatagateway.org/dataset/fddcb382-3051-4394-8436-b92295f14259). US investigators are encouraged to coordinate data requests through the Coronavirus Pandemic Epidemiology Consortium (https://www.monganinstitute.org/cope-consortium).

https://web.www.healthdatagateway.org/dataset/fddcb382-3051-4394-8436-b92295f14259

## Ethics

Ethics has been approved by KCL Ethics Committee REMAS ID 18210, review reference LRS-19/20-18210 and all participants provided consent.

## Author Contributions

MSG, CHS, ATC, JW, TDS, CJS, SO contributed to study concept and design. CHS, AM, BM, DAD, LHN, LP, SS, CH, JCP, The COVID-19 Genomics UK (COG-UK) consortium, ATC, JW, TDS, CJS, SO contributed to acquisition of data. MSG, CHS, AM, TV, contributed to data analysis and have verified the underlying data. MSG, CHS contributed to initial drafting of the manuscript. MGS, CHS, CJS and SO were responsible for the decision to submit the manuscript. All authors contributed to interpretation of data and critical revision of the manuscript. ATC, CJS, TDS, SO contributed to study supervision.

## Acknowledgements

ZOE Global provided in kind support for all aspects of building, running and supporting the app and service to all users worldwide. COG-UK is supported by funding from the Medical Research Council (MRC) part of UK Research & Innovation (UKRI), the National Institute of Health Research (NIHR) and Genome Research Limited, operating as the Wellcome Sanger Institute. Support for this study was provided by the NIHR-funded Biomedical Research Centre based at GSTT NHS Foundation Trust. Investigators also received support from the Wellcome Trust (212904/Z/18/Z, WT203148/Z/16/Z), the MRC/BHF (MR/M016560/1), Alzheimer’s Society, EU, NIHR, CDRF, and the NIHR-funded BioResource, Clinical Research Facility and BRC based at GSTT NHS Foundation Trust in partnership with KCL, the UK Research and Innovation London Medical Imaging & Artificial Intelligence Centre for Value Based Healthcare, the Wellcome Flagship Programme (WT213038/Z/18/Z), the Chronic Disease Research Foundation, and DHSC. ATC was supported in this work through a Stuart and Suzanne Steele MGH Research Scholar Award. The Massachusetts Consortium on Pathogen Readiness (MassCPR) and Mark and Lisa Schwartz supported MGH investigators (DAD, LHN, ATC).

## Supplementary material

**Supplementary Figure 1.**
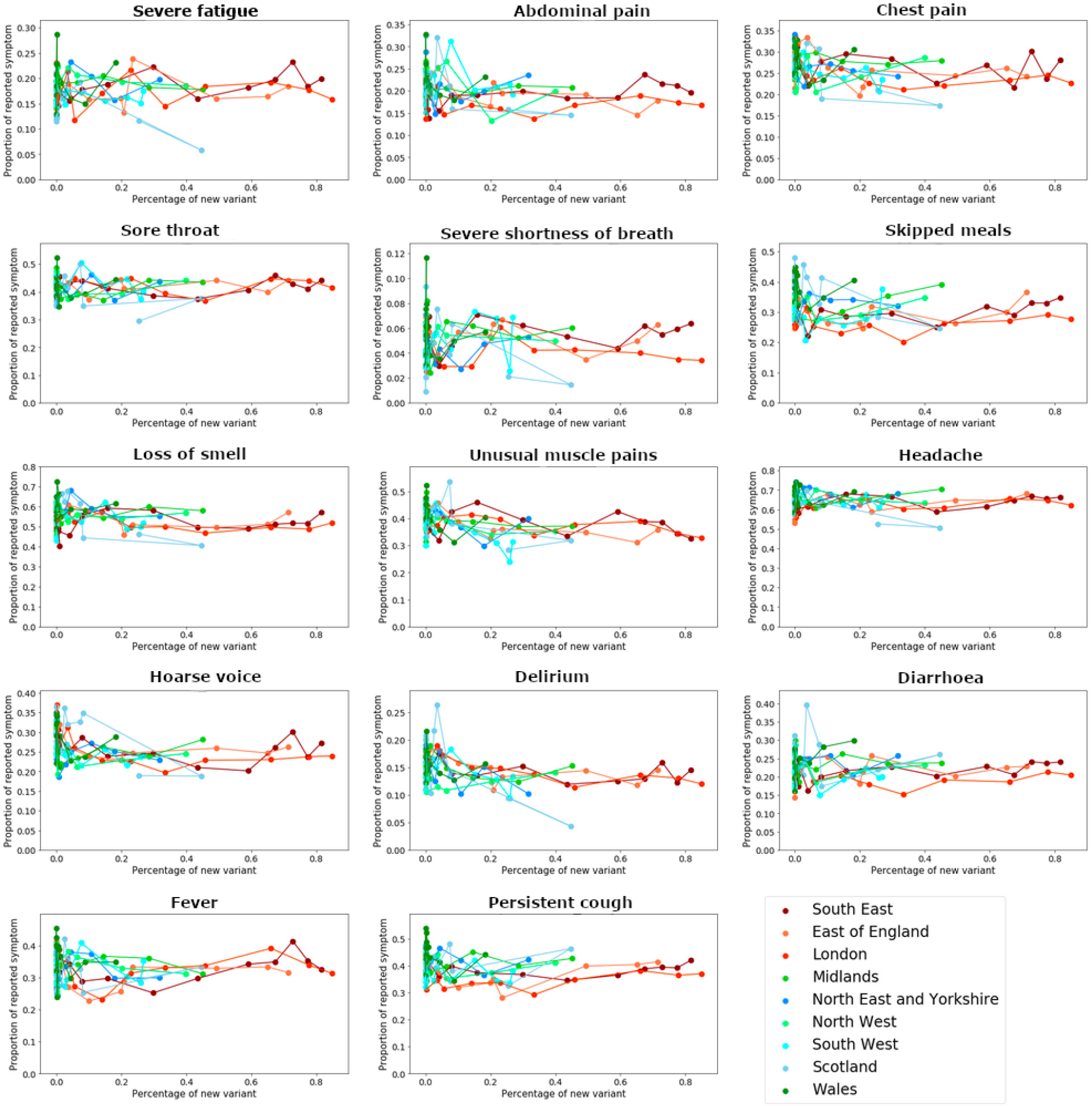
Regional plots of the frequency of reporting of symptoms over time for each reported symptom, against the proportion of B.1.1.7.. Drop in fever reporting in early November was caused by a change in the question wording; this wording was subsequently reverted a week later.

**Supplementary Figure 2.**
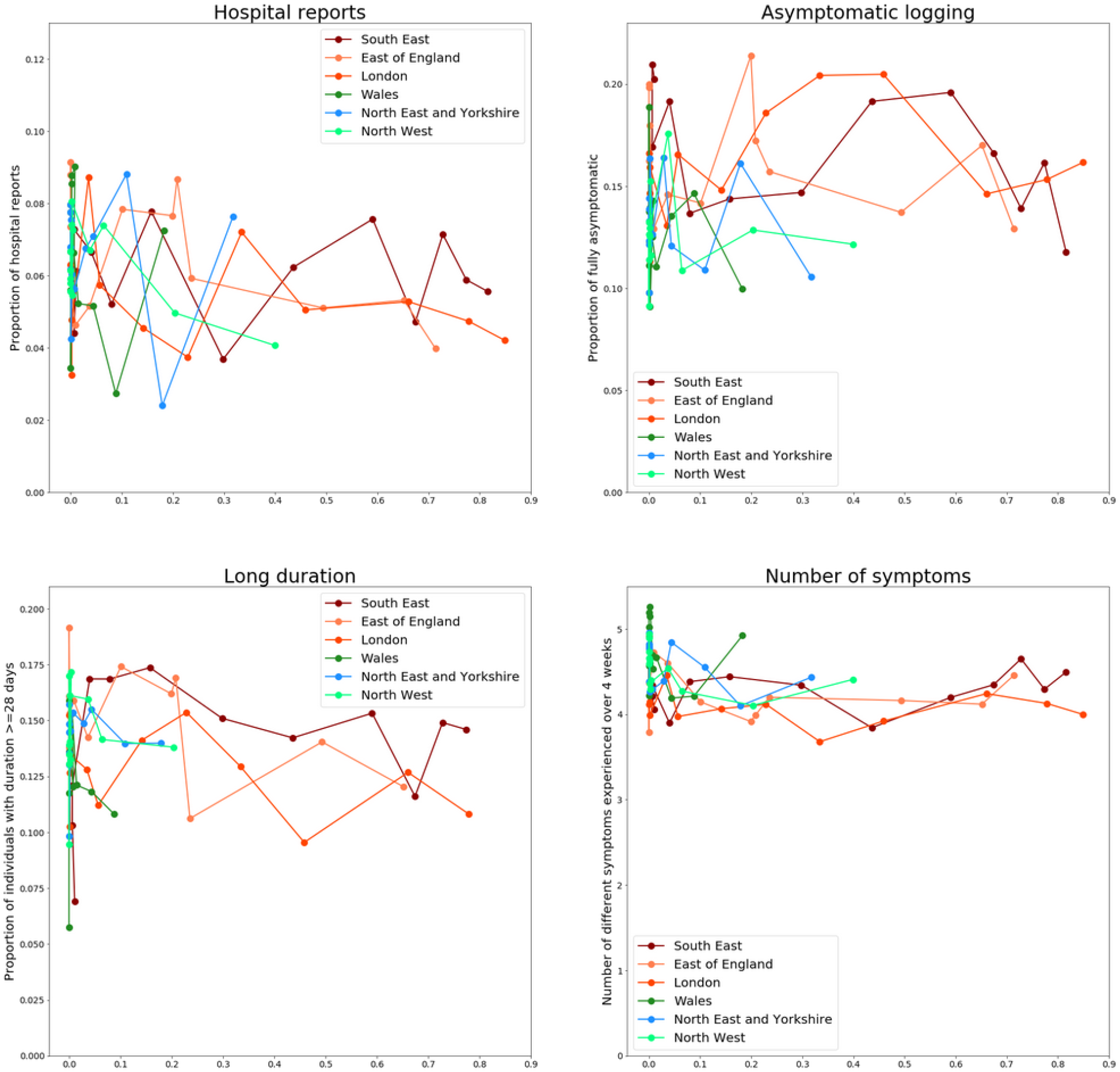
Regional plot of hospitalisation reports, proportion of asymptomatic positives, instances of long symptom duration and the total number of different experienced symptoms against proportion of B.1.1.7. For the study of long symptom duration, tests are only considered up to 21 December, and symptom reports up to 18 January 2021 to limit right censoring effects. Only symptomatic individuals for which duration can be ascertained are included.

**Supplementary Figure 3.**
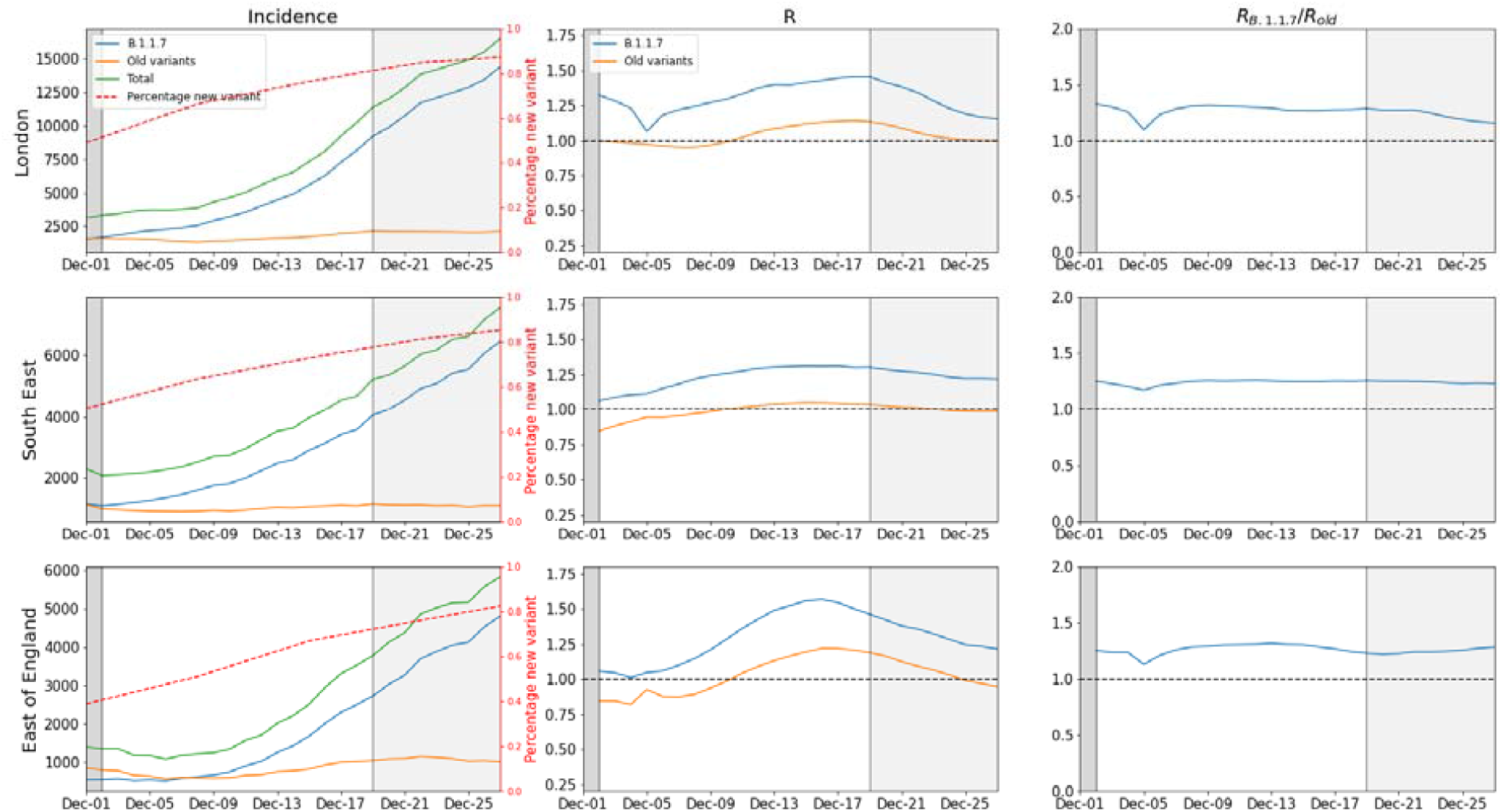
Incidence and R(t) for the old and new variants, along with the ratio between these R values, for the three regions in England with the largest proportion of B.1.1.7, using SGTF data. Dark grey regions indicate national lockdowns, light grey shaded the period where London and much of the South East and East of England were placed in Tier 4 restrictions.

**Supplementary Figure 4:**
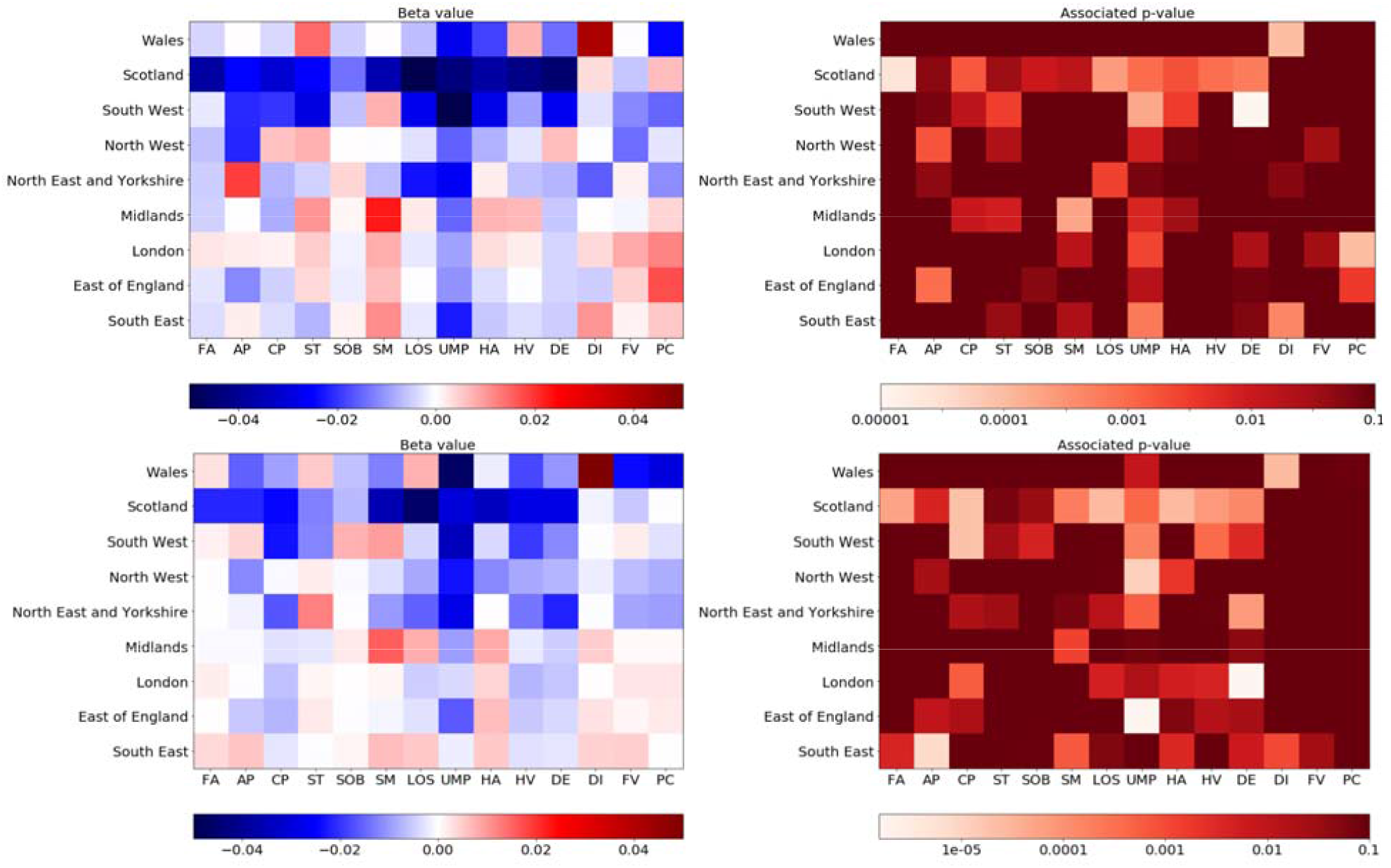
Colour plot of beta values and associated p-values for each region and symptom when investigating association between symptom report (in a 4 week window around the test) and proportion of variant B.1.1.7. Top row shows results for uncorrected model, and the bottom row shows results for the model corrected for age, sex, temperature and humidity. Note that the p-values are capped at 0.1. Beta-values are presented for an increase of 0.1 in the proportion of variant B.1.1.7. Key: FA - fatigue, AP - abdominal pain, CP - chest pain, ST - sore throat, SOB - shortness of breath, SM - skipped meals, LOS - loss of smell, UMP - unusual muscle pains, HA - headache, HV - hoarse voice, DE - delirium, DI - diarrhoea, FV - fever, PC - persistent cough

**Supplementary Figure 5:**
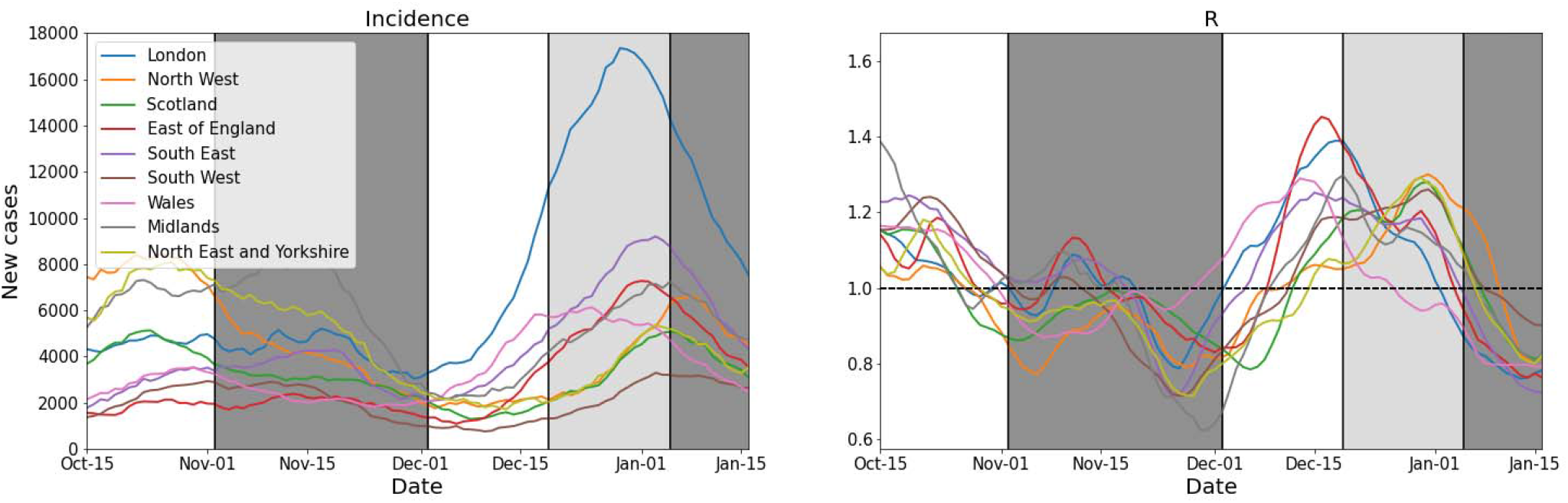
Total incidence and R(t) for all regions, extended to capture the third national lockdown beginning 5 January 2021. Dark grey regions indicate national lockdowns, light grey indicate the period where London and much of the South East and East of England were placed in Tier 4 restrictions.

**Supplementary Table 1:**
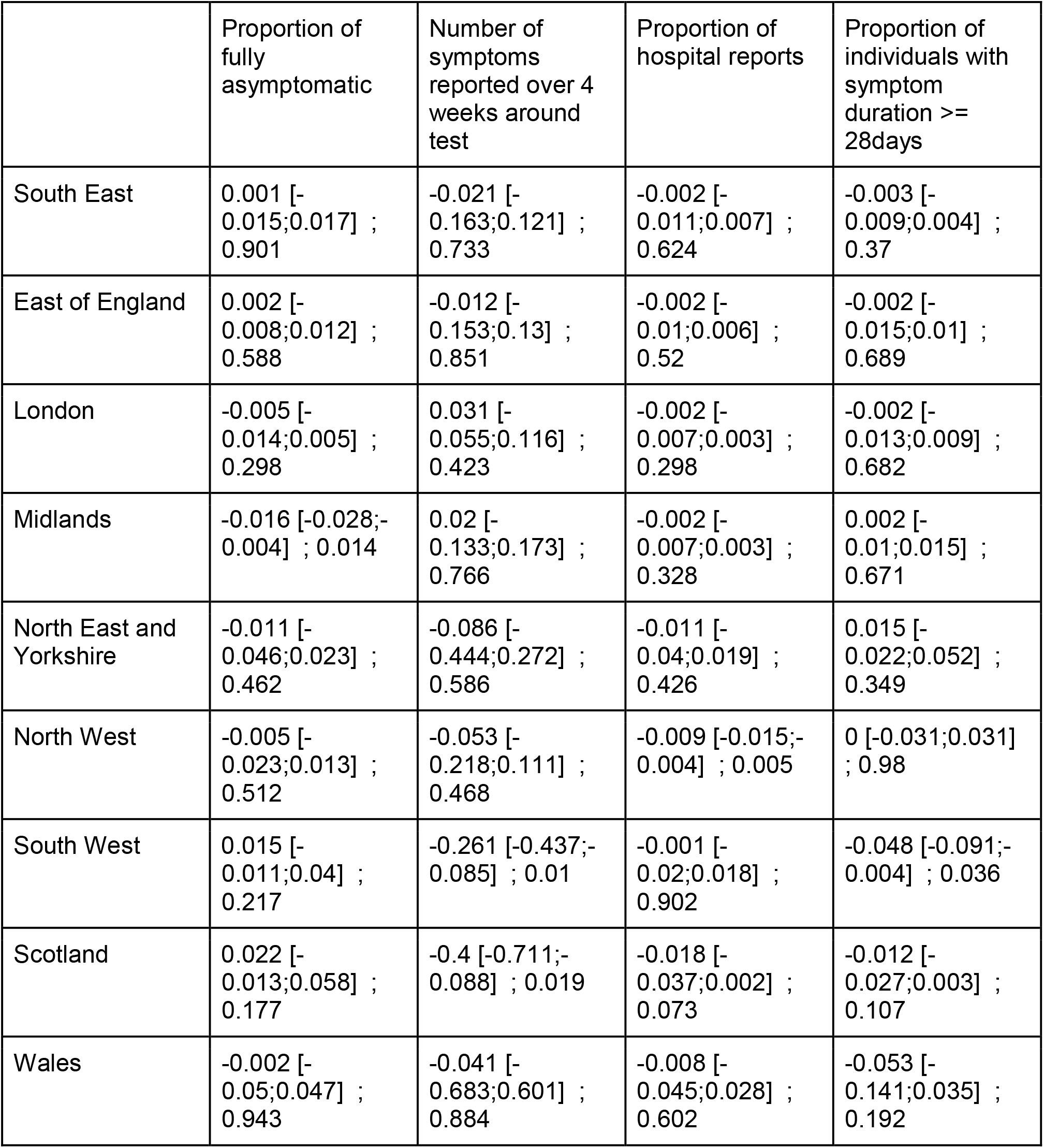
Beta coefficient of the variant proportion when evaluating association with number of reported symptoms, asymptomatic rate, proportion of hospital report and proportion of individuals with duration >28 days (among symptomatic) across the different regions when correcting for age, sex, temperature and humidity. All values are presented for an increase in 0.1 in the proportion of variant B.1.1.7. All results are presented in the form mean [CI]; p-value

| | Proportion of fully asymptomatic | Number of symptoms reported over 4 weeks around test | Proportion of hospital reports | Proportion of individuals with symptom duration ≥ 28days |
| --- | --- | --- | --- | --- |
| South East | 0.001 [-0.015;0.017] ; 0.901 | -0.021 [-0.163;0.121] ; 0.733 | -0.002 [-0.011;0.007] ; 0.624 | -0.003 [-0.009;0.004] ; 0.37 |
| East of England | 0.002 [-0.008;0.012] ; 0.588 | -0.012 [-0.153;0.13] ; 0.851 | -0.002 [-0.01;0.006] ; 0.52 | -0.002 [-0.015;0.01] ; 0.689 |
| London | -0.005 [-0.014;0.005] ; 0.298 | 0.031 [-0.055;0.116] ; 0.423 | -0.002 [-0.007;0.003] ; 0.298 | -0.002 [-0.013;0.009] ; 0.682 |
| Midlands | -0.016 [-0.028;-0.004] ; 0.014 | 0.02 [-0.133;0.173] ; 0.766 | -0.002 [-0.007;0.003] ; 0.328 | 0.002 [-0.01;0.015] ; 0.671 |
| North East and Yorkshire | -0.011 [-0.046;0.023] ; 0.462 | -0.086 [-0.444;0.272] ; 0.586 | -0.011 [-0.04;0.019] ; 0.426 | 0.015 [-0.022;0.052] ; 0.349 |
| North West | -0.005 [-0.023;0.013] ; 0.512 | -0.053 [-0.218;0.111] ; 0.468 | -0.009 [-0.015;-0.004] ; 0.005 | 0 [-0.031;0.031] ; 0.98 |
| South West | 0.015 [-0.011;0.04] ; 0.217 | -0.261 [-0.437;-0.085] ; 0.01 | -0.001 [-0.02;0.018] ; 0.902 | -0.048 [-0.091;-0.004] ; 0.036 |
| Scotland | 0.022 [-0.013;0.058] ; 0.177 | -0.4 [-0.711;-0.088] ; 0.019 | -0.018 [-0.037;0.002] ; 0.073 | -0.012 [-0.027;0.003] ; 0.107 |
| Wales | -0.002 [-0.05;0.047] ; 0.943 | -0.041 [-0.683;0.601] ; 0.884 | -0.008 [-0.045;0.028] ; 0.602 | -0.053 [-0.141;0.035] ; 0.192 |

**Supplementary Table 2:**
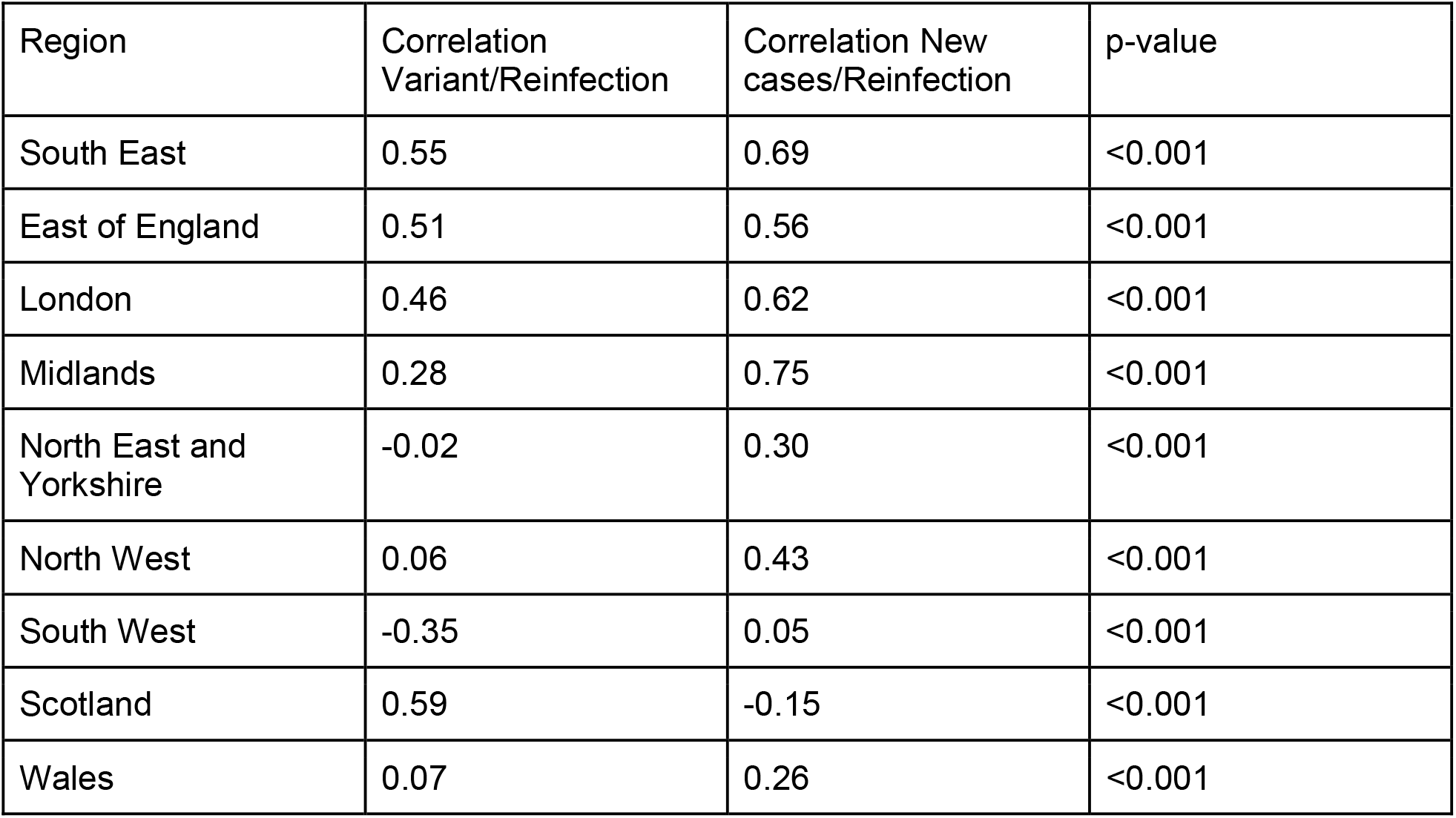
Comparison of regional correlation over time between proportion of B.1.1.7 and number of possible reinfections and between new reported cases and number of possible reinfections. Medians over 100 bootstrapped samples are calculated for each and compared using a Mann-Whitney U test.

| Region | Correlation Variant/Reinfection | Correlation New cases/Reinfection | p-value |
| --- | --- | --- | --- |
| South East | 0.55 | 0.69 | <0.001 |
| East of England | 0.51 | 0.56 | <0.001 |
| London | 0.46 | 0.62 | <0.001 |
| Midlands | 0.28 | 0.75 | <0.001 |
| North East and Yorkshire | -0.02 | 0.30 | <0.001 |
| North West | 0.06 | 0.43 | <0.001 |
| South West | -0.35 | 0.05 | <0.001 |
| Scotland | 0.59 | -0.15 | <0.001 |
| Wales | 0.07 | 0.26 | <0.001 |

COG-UK authorship list

**Funding acquisition, Leadership and supervision, Metadata curation, Project administration, Samples and logistics, Sequencing and analysis, Software and analysis tools, and Visualisation:** Samuel C Robson ^13^.

**Funding acquisition, Leadership and supervision, Metadata curation, Project administration, Samples and logistics, Sequencing and analysis, and Software and analysis tools:**

Nicholas J Loman ^41^ and Thomas R Connor ^10, 69^.

**Leadership and supervision, Metadata curation, Project administration, Samples and logistics, Sequencing and analysis, Software and analysis tools, and Visualisation:**

Tanya Golubchik ^5^.

**Funding acquisition, Metadata curation, Samples and logistics, Sequencing and analysis, Software and analysis tools, and Visualisation:**

Rocio T Martinez Nunez ^42^.

**Funding acquisition, Leadership and supervision, Metadata curation, Project administration, and Samples and logistics:**

Catherine Ludden ^88^.

**Funding acquisition, Leadership and supervision, Metadata curation, Samples and logistics, and Sequencing and analysis:**

Sally Corden ^69^.

**Funding acquisition, Leadership and supervision, Project administration, Samples and logistics, and Sequencing and analysis:**

Ian Johnston ^99^ and David Bonsall ^5^.

**Funding acquisition, Leadership and supervision, Sequencing and analysis, Software and analysis tools, and Visualisation:**

Colin P Smith ^87^ and Ali R Awan ^28^.

**Funding acquisition, Samples and logistics, Sequencing and analysis, Software and analysis tools, and Visualisation:**

Giselda Bucca ^87^.

**Leadership and supervision, Metadata curation, Project administration, Samples and logistics, and Sequencing and analysis:**

M. Estee Torok ^22, 101^.

**Leadership and supervision, Metadata curation, Project administration, Samples and logistics, and Visualisation:**

Kordo Saeed ^81, 110^ and Jacqui A Prieto ^83, 109^.

**Leadership and supervision, Metadata curation, Project administration, Sequencing and analysis, and Software and analysis tools:**

David K Jackson ^99^.

**Metadata curation, Project administration, Samples and logistics, Sequencing and analysis, and Software and analysis tools:**

William L Hamilton ^22^.

**Metadata curation, Project administration, Samples and logistics, Sequencing and analysis, and Visualisation:**

Luke B Snell ^11^.

**Funding acquisition, Leadership and supervision, Metadata curation, and Samples and logistics:**

Catherine Moore ^69^.

**Funding acquisition, Leadership and supervision, Project administration, and Samples and logistics:**

Ewan M Harrison ^99, 88^.

**Leadership and supervision, Metadata curation, Project administration, and Samples and logistics:**

Sonia Goncalves ^99^.

**Leadership and supervision, Metadata curation, Samples and logistics, and Sequencing and analysis:**

Ian G Goodfellow ^24^, Derek J Fairley ^3, 72^, Matthew W Loose ^18^ and Joanne Watkins ^69^.

**Leadership and supervision, Metadata curation, Samples and logistics, and Software and analysis tools:**

Rich Livett ^99^.

**Leadership and supervision, Metadata curation, Samples and logistics, and Visualisation:**

Samuel Moses ^25, 106^.

**Leadership and supervision, Metadata curation, Sequencing and analysis, and Software and analysis tools:**

Roberto Amato ^99^, Sam Nicholls ^41^ and Matthew Bull ^69^.

**Leadership and supervision, Project administration, Samples and logistics, and Sequencing and analysis:**

Darren L Smith ^37, 58, 105^.

**Leadership and supervision, Sequencing and analysis, Software and analysis tools, and Visualisation:**

Jeff Barrett ^99^, David M Aanensen ^14, 114^.

**Metadata curation, Project administration, Samples and logistics, and Sequencing and analysis:**

Martin D Curran ^65^, Surendra Parmar ^65^, Dinesh Aggarwal ^95, 99, 64^ and James G Shepherd ^48^.

**Metadata curation, Project administration, Sequencing and analysis, and Software and analysis tools:**

Matthew D Parker ^93^.

**Metadata curation, Samples and logistics, Sequencing and analysis, and Visualisation:**

Sharon Glaysher ^61^.

**Metadata curation, Sequencing and analysis, Software and analysis tools, and Visualisation:**

Matthew Bashton ^37, 58^, Anthony P Underwood ^14, 114^, Nicole Pacchiarini ^69^ and Katie F Loveson ^77^.

**Project administration, Sequencing and analysis, Software and analysis tools, and Visualisation:**

Alessandro M Carabelli ^88^.

**Funding acquisition, Leadership and supervision, and Metadata curation:**

Kate E Templeton ^53, 90^.

**Funding acquisition, Leadership and supervision, and Project administration:**

Cordelia F Langford ^99^, John Sillitoe ^99^, Thushan I de Silva ^93^ and Dennis Wang ^93^.

**Funding acquisition, Leadership and supervision, and Sequencing and analysis:**

Dominic Kwiatkowski ^99, 107^, **And**rew Rambaut ^90^, Justin O’Grady ^70, 89^ and Simon Cottrell ^69^.

**Leadership and supervision, Metadata curation, and Sequencing and analysis:**

Matthew T.G. Holden ^68^ and Emma C Thomson ^48^.

**Leadership and supervision, Project administration, and Samples and logistics:**

Husam Osman ^64, 36^, Monique Andersson ^59^, Anoop J Chauhan ^61^ and Mohammed O Hassan-Ibrahim ^6^.

**Leadership and supervision, Project administration, and Sequencing and analysis:**

Mara Lawniczak ^99^.

**Leadership and supervision, Samples and logistics, and Sequencing and analysis:**

Ravi Kumar Gupta ^88, 113^, Alex Alderton ^99^, Meera Chand ^66^, Chrystala Constantinidou ^94^, Meera Unnikrishnan ^94^, Alistair C Darby ^92^, Julian A Hiscox ^92^ and Steve Paterson ^92^.

**Leadership and supervision, Sequencing and analysis, and Software and analysis tools:**

Inigo Martincorena ^99^, David L Robertson ^48^, Erik M Volz ^39^, Andrew J Page ^70^ and Oliver G Pybus ^23^.

**Leadership and supervision, Sequencing and analysis, and Visualisation:**

Andrew R Bassett ^99^.

**Metadata curation, Project administration, and Samples and logistics:**

Cristina V Ariani ^99^, Michael H Spencer Chapman ^99, 88^, Kathy K Li ^48^, Rajiv N Shah ^48^, Natasha G Jesudason ^48^ and Yusri Taha ^50^.

**Metadata curation, Project administration, and Sequencing and analysis:**

Martin P McHugh ^53^ and Rebecca Dewar ^53^.

**Metadata curation, Samples and logistics, and Sequencing and analysis:**

Aminu S Jahun ^24^, Claire McMurray ^41^, Sarojini Pandey ^84^, James P McKenna ^3^, Andrew Nelson ^58, 105^, Gregory R Young ^37, 58^, Clare M McCann ^58, 105^ and Scott Elliott ^61^.

**Metadata curation, Samples and logistics, and Visualisation:**

Hannah Lowe ^25^.

**Metadata curation, Sequencing and analysis, and Software and analysis tools:**

Ben Temperton ^91^, Sunando Roy ^82^, Anna Price ^10^, Sara Rey ^69^ and Matthew Wyles ^93^.

**Metadata curation, Sequencing and analysis, and Visualisation:**

Stefan Rooke ^90^ and Sharif Shaaban ^68^.

**Project administration, Samples and logistics, Sequencing and analysis:**

Mariateresa de Cesare ^98^.

**Project administration, Samples and logistics, and Software and analysis tools:**

Laura Letchford ^99^.

**Project administration, Samples and logistics, and Visualisation:**

Siona Silveira ^81^, Emanuela Pelosi ^81^ and Eleri Wilson-Davies ^81^.

**Samples and logistics, Sequencing and analysis, and Software and analysis tools:**

Myra Hosmillo ^24^.

**Sequencing and analysis, Software and analysis tools, and Visualisation:**

Áine O’Toole ^90^, Andrew R Hesketh ^87^, Richard Stark ^94^, Louis du Plessis ^23^, Chris Ruis ^88^, Helen Adams ^4^ and Yann Bourgeois ^76^.

**Funding acquisition, and Leadership and supervision:**

Stephen L Michell ^91^, Dimitris Grammatopoulos^84, 112^, Jonathan Edgeworth ^12^, Judith Breuer ^30, 82^, John A Todd ^98^ and Christophe Fraser ^5^.

**Funding acquisition, and Project administration:**

David Buck ^98^ and Michaela John ^9^.

**Leadership and supervision, and Metadata curation:**

Gemma L Kay ^70^.

**Leadership and supervision, and Project administration:**

Steve Palmer ^99^, Sharon J Peacock ^88, 64^ and David Heyburn ^69^.

**Leadership and supervision, and Samples and logistics:**

Danni Weldon ^99^, Esther Robinson ^64, 36^, Alan McNally ^41, 86^, Peter Muir ^64^, Ian B Vipond ^64^, John BoYes ^29^, Venkat Sivaprakasam ^46^, Tranprit Salluja ^75^, Samir Dervisevic ^54^ and Emma J Meader ^54^.

**Leadership and supervision, and Sequencing and analysis:**

Naomi R Park ^99^, Karen Oliver ^99^, Aaron R Jeffries ^91^, Sascha Ott ^94^, Ana da Silva Filipe ^48^, David A Simpson ^72^ and Chris Williams ^69^.

**Leadership and supervision, and Visualisation:**

Jane AH Masoli ^73, 91^.

**Metadata curation, and Samples and logistics:**

Bridget A Knight ^73, 91^, Christopher R Jones ^73, 91^, Cherian Koshy ^1^, Amy Ash ^1^, Anna Casey ^71^, Andrew Bosworth ^64, 36^, Liz Ratcliffe ^71^, Li Xu-McCrae ^36^, Hannah M Pymont ^64^, Stephanie Hutchings ^64^, Lisa Berry ^84^, Katie Jones ^84^, Fenella Halstead ^46^, Thomas Davis ^21^, Christopher Holmes ^16^, Miren Iturriza-Gomara ^92^, Anita O Lucaci ^92^, Paul Anthony Randell ^38, 104^, Alison Cox ^38, 104^, Pinglawathee Madona ^38, 104^, Kathryn Ann Harris ^30^, Julianne Rose Brown ^30^, Tabitha W Mahungu ^74^, Dianne Irish-Tavares ^74^, Tanzina Haque ^74^, Jennifer Hart ^74^, Eric Witele ^74^, Melisa Louise Fenton ^75^, Steven Liggett ^79^, Clive Graham ^56^, Emma Swindells ^57^, Jennifer Collins ^50^, Gary Eltringham ^50^, Sharon Campbell ^17^, Patrick C McClure ^97^, Gemma Clark ^15^, Tim J Sloan ^60^, Carl Jones ^15^ and Jessica Lynch ^2, 111^.

**Metadata curation, and Sequencing and analysis:**

Ben Warne ^8^, Steven Leonard ^99^, Jillian Durham ^99^, Thomas Williams ^90^, Sam T Haldenby ^92^, Nathaniel Storey ^30^, Nabil-Fareed Alikhan ^70^, Nadine Holmes ^18^, Christopher Moore ^18^, Matthew Carlile ^18^, Malorie Perry ^69^, Noel Craine ^69^, Ronan A Lyons ^80^, Angela H Beckett ^13^, Salman Goudarzi ^77^, Christopher Fearn ^77^, Kate Cook ^77^, Hannah Dent ^77^ and Hannah Paul ^77^.

**Metadata curation, and Software and analysis tools:**

Robert Davies ^99^.

**Project administration, and Samples and logistics:**

Beth Blane ^88^, Sophia T Girgis ^88^, Mathew A Beale ^99^, Katherine L Bellis ^99, 88^, Matthew J Dorman ^99^, Eleanor Drury ^99^, Leanne Kane ^99^, Sally Kay ^99^, Samantha McGuigan ^99^, Rachel Nelson ^99^, Liam Prestwood ^99^, Shavanthi Rajatileka ^99^, Rahul Batra ^12^, Rachel J Williams ^82^, Mark Kristiansen ^82^, Angie Green ^98^, Anita Justice ^59^, Adhyana I.K Mahanama ^81, 102^ and Buddhini Samaraweera ^81, 102^.

**Project administration, and Sequencing and analysis:**

Nazreen F Hadjirin ^88^ and Joshua Quick ^41^.

**Project administration, and Software and analysis tools:**

Radoslaw Poplawski ^41^.

**Samples and logistics, and Sequencing and analysis:**

Leanne M Kermack ^88^, Nicola Reynolds ^7^, Grant Hall ^24^, Yasmin Chaudhry ^24^, Malte L Pinckert ^24^, Iliana Georgana ^24^, Robin J Moll ^99^, Alicia Thornton ^66^, Richard Myers ^66^, Joanne Stockton ^41^, Charlotte A Williams ^82^, Wen C Yew ^58^, Alexander J Trotter ^70^, Amy Trebes ^98^, George MacIntyre-Cockett ^98^, Alec Birchley ^69^, Alexander Adams ^69^, Amy Plimmer ^69^, Bree Gatica-Wilcox ^69^, Caoimhe McKerr ^69^, Ember Hilvers ^69^, Hannah Jones ^69^, Hibo Asad ^69^, Jason Coombes ^69^, Johnathan M Evans ^69^, Laia Fina ^69^, Lauren Gilbert ^69^, Lee Graham ^69^, Michelle Cronin ^69^, Sara Kumziene-SummerhaYes ^69^, Sarah Taylor ^69^, Sophie Jones ^69^, Danielle C Groves ^93^, Peijun Zhang ^93^, Marta Gallis ^93^ and Stavroula F Louka ^93^.

**Samples and logistics, and Software and analysis tools:**

Igor Starinskij ^48^.

**Sequencing and analysis, and Software and analysis tools:**

Chris J Illingworth ^47^, Chris Jackson ^47^, Marina Gourtovaia ^99^, Gerry Tonkin-Hill ^99^, Kevin Lewis ^99^, Jaime M Tovar-Corona ^99^, Keith James ^99^, Laura Baxter ^94^, Mohammad T. Alam ^94^, Richard J Orton ^48^, Joseph Hughes ^48^, Sreenu Vattipally ^48^, Manon Ragonnet-Cronin ^39^, Fabricia F. Nascimento ^39^, David Jorgensen ^39^, Olivia Boyd ^39^, Lily Geidelberg ^39^, Alex E Zarebski ^23^, Jayna Raghwani ^23^, Moritz UG Kraemer ^23^, Joel Southgate ^10, 69^, Benjamin B Lindsey ^93^ and Timothy M Freeman ^93^.

**Software and analysis tools, and Visualisation:**

Jon-Paul Keatley ^99^, Joshua B Singer ^48^, Leonardo de Oliveira Martins ^70^, Corin A Yeats ^14^, Khalil Abudahab ^14, 114^, Ben EW Taylor ^14, 114^ and Mirko Menegazzo ^14^.

**Leadership and supervision:**

John Danesh ^99^, Wendy Hogsden ^46^, Sahar Eldirdiri ^21^, Anita Kenyon ^21^, Jenifer Mason ^43^, Trevor I Robinson ^43^, Alison Holmes ^38, 103^, James Price ^38, 103^, John A Hartley ^82^, Tanya Curran ^3^, Alison E Mather ^70^, Giri Shankar ^69^, Rachel Jones ^69^, Robin Howe ^69^ and Sian Morgan ^9^.

**Metadata curation:**

Elizabeth Wastenge ^53^, Michael R Chapman ^34, 88, 99^, Siddharth Mookerjee ^38, 103^, Rachael Stanley ^54^, Wendy Smith ^15^, Timothy Peto ^59^, David Eyre ^59^, Derrick Crook ^59^, Gabrielle Vernet ^33^, Christine Kitchen ^10^, Huw Gulliver ^10^, Ian Merrick ^10^, Martyn Guest ^10^, Robert Munn ^10^, Declan T Bradley ^63, 72^ and Tim Wyatt ^63^.

**Project administration:**

Charlotte Beaver ^99^, Luke Foulser ^99^, Sophie Palmer ^88^, Carol M Churcher ^88^, Ellena Brooks ^88^, Kim S Smith ^88^, Katerina Galai ^88^, Georgina M McManus ^88^, Frances Bolt ^38, 103^, Francesc Coll ^19^, Lizzie Meadows ^70^, Stephen W Attwood ^23^, Alisha Davies ^69^, Elen De Lacy ^69^, Fatima Downing ^69^, Sue Edwards ^69^, Garry P Scarlett ^76^, Sarah Jeremiah ^83^ and Nikki Smith ^93^.

**Samples and logistics:**

Danielle Leek ^88^, Sushmita Sridhar ^88, 99^, Sally Forrest ^88^, Claire Cormie ^88^, Harmeet K Gill ^88^, Joana Dias ^88^, Ellen E Higginson ^88^, Mailis Maes ^88^, Jamie Young ^88^, Michelle Wantoch ^7^, Sanger Covid Team (www.sanger.ac.uk/covid-team) ^99^, Dorota Jamrozy ^99^, Stephanie Lo ^99^, Minal Patel ^99^, Verity Hill ^90^, Claire M Bewshea ^91^, Sian Ellard ^73, 91^, Cressida Auckland ^73^, Ian Harrison ^66^, Chloe Bishop ^66^, Vicki Chalker ^66^, Alex Richter ^85^, Andrew Beggs ^85^, Angus Best ^86^, Benita Percival ^86^, Jeremy Mirza ^86^, Oliver Megram ^86^, Megan Mayhew ^86^, Liam Crawford ^86^, Fiona Ashcroft ^86^, Emma Moles-Garcia ^86^, Nicola Cumley ^86^, Richard Hopes ^64^, Patawee Asamaphan ^48^, Marc O Niebel ^48^, Rory N Gunson ^100^, Amanda Bradley ^52^, Alasdair Maclean ^52^, Guy Mollett ^52^, Rachel Blacow ^52^, Paul Bird ^16^, Thomas Helmer ^16^, Karlie Fallon ^16^, Julian Tang ^16^, Antony D Hale ^49^, Louissa R Macfarlane-Smith ^49^, Katherine L Harper ^49^, Holli Carden ^49^, Nicholas W Machin ^45, 64^, Kathryn A Jackson ^92^, Shazaad S Y Ahmad ^45, 64^, Ryan P George ^45^, Lance Turtle ^92^, Elaine O’Toole ^43^, Joanne Watts ^43^, Cassie Breen ^43^, Angela Cowell ^43^, Adela Alcolea-Medina ^32, 96^, Themoula Charalampous ^12, 42^, Amita Patel ^11^, Lisa J Levett ^35^, Judith Heaney ^35^, Aileen Rowan ^39^, Graham P Taylor ^39^, Divya Shah ^30^, Laura Atkinson ^30^, Jack CD Lee ^30^, Adam P Westhorpe ^82^, Riaz Jannoo ^82^, Helen L Lowe ^82^, Angeliki Karamani ^82^, Leah Ensell ^82^, Wendy Chatterton ^35^, Monika Pusok ^35^, Ashok Dadrah ^75^, Amanda Symmonds ^75^, Graciela Sluga ^44^, Zoltan Molnar ^72^, Paul Baker ^79^, Stephen Bonner ^79^, Sarah Essex ^79^, Edward Barton ^56^, Debra Padgett ^56^, Garren Scott ^56^, Jane Greenaway ^57^, Brendan AI Payne ^50^, Shirelle Burton-Fanning ^50^, Sheila Waugh ^50^, Veena Raviprakash ^17^, Nicola Sheriff ^17^, Victoria Blakey ^17^, Lesley-Anne Williams ^17^, Jonathan Moore ^27^, Susanne Stonehouse ^27^, Louise Smith ^55^, Rose K Davidson ^89^, Luke Bedford ^26^, Lindsay Coupland ^54^, Victoria Wright ^18^, Joseph G Chappell ^97^, Theocharis Tsoleridis ^97^, Jonathan Ball ^97^, Manjinder Khakh ^15^, Vicki M Fleming ^15^, Michelle M Lister ^15^, Hannah C Howson-Wells ^15^, Louise Berry ^15^, Tim Boswell ^15^, Amelia Joseph ^15^, Iona Willingham ^15^, Nichola Duckworth ^60^, Sarah Walsh ^60^, Emma Wise ^2, 111^, Nathan Moore ^2, 111^, Matilde Mori ^2, 108, 111^, Nick Cortes ^2, 111^, Stephen Kidd ^2, 111^, Rebecca Williams ^33^, Laura Gifford ^69^, Kelly Bicknell ^61^, Sarah Wyllie ^61^, Allyson Lloyd ^61^, Robert Impey ^61^, Cassandra S Malone ^6^, Benjamin J Cogger ^6^, Nick Levene ^62^, Lynn Monaghan ^62^, Alexander J Keeley ^93^, David G Partridge ^78, 93^, Mohammad Raza ^78, 93^, Cariad Evans ^78, 93^ and Kate Johnson ^78, 93^.

**Sequencing and analysis:**

Emma Betteridge ^99^, Ben W Farr ^99^, Scott Goodwin ^99^, Michael A Quail ^99^, Carol Scott ^99^, Lesley Shirley ^99^, Scott AJ Thurston ^99^, Diana Rajan ^99^, Iraad F Bronner ^99^, Louise Aigrain ^99^, Nicholas M Redshaw ^99^, Stefanie V Lensing ^99^, Shane McCarthy ^99^, Alex Makunin ^99^, Carlos E Balcazar ^90^, Michael D Gallagher ^90^, Kathleen A Williamson ^90^, Thomas D Stanton ^90^, Michelle L Michelsen ^91^, Joanna Warwick-Dugdale ^91^, Robin Manley ^91^, Audrey Farbos ^91^, James W Harrison ^91^, Christine M Sambles ^91^, David J Studholme ^91^, Angie Lackenby ^66^, Tamyo Mbisa ^66^, Steven Platt ^66^, Shahjahan Miah ^66^, David Bibby ^66^, Carmen Manso ^66^, Jonathan Hubb ^66^, Gavin Dabrera ^66^, Mary Ramsay ^66^, Daniel Bradshaw ^66^, Ulf Schaefer ^66^, Natalie Groves ^66^, Eileen Gallagher ^66^, David Lee ^66^, David Williams ^66^, Nicholas Ellaby ^66^, Hassan Hartman ^66^, Nikos Manesis ^66^, Vineet Patel ^66^, Juan Ledesma ^67^, Katherine A Twohig ^67^, Elias Allara ^64, 88^, Clare Pearson ^64, 88^, Jeffrey K. J. Cheng ^94^, Hannah E. Bridgewater ^94^, Lucy R. Frost ^94^, Grace Taylor-Joyce ^94^, Paul E Brown ^94^, Lily Tong ^48^, Alice Broos ^48^, Daniel Mair ^48^, Jenna Nichols ^48^, Stephen N Carmichael ^48^, Katherine L Smollett ^40^, Kyriaki Nomikou ^48^, Elihu Aranday-Cortes ^48^, Natasha Johnson ^48^, Seema Nickbakhsh ^48, 68^, Edith E Vamos ^92^, Margaret Hughes ^92^, Lucille Rainbow ^92^, Richard Eccles ^92^, Charlotte

Nelson ^92^, Mark Whitehead ^92^, Richard Gregory ^92^, Matthew Gemmell ^92^, Claudia Wierzbicki ^92^, Hermione J Webster ^92^, Chloe L Fisher ^28^, Adrian W Signell ^20^, Gilberto Betancor ^20^, Harry D Wilson ^20^, Gaia Nebbia ^12^, Flavia Flaviani ^31^, Alberto C Cerda ^96^, Tammy V Merrill ^96^, Rebekah E Wilson ^96^, Marius Cotic ^82^, Nadua Bayzid ^82^, Thomas Thompson ^72^, Erwan Acheson ^72^, Steven Rushton ^51^, Sarah O’Brien ^51^, David J Baker ^70^, Steven Rudder ^70^, Alp Aydin ^70^, Fei Sang ^18^, Johnny Debebe ^18^, Sarah Francois ^23^, Tetyana I Vasylyeva ^23^, Marina Escalera Zamudio ^23^, Bernardo Gutierrez ^23^, Angela Marchbank ^10^, Joshua Maksimovic ^9^, Karla Spellman ^9^, Kathryn McCluggage ^9^, Mari Morgan ^69^, Robert Beer ^9^, Safiah Afifi ^9^, Trudy Workman ^10^, William Fuller ^10^, Catherine Bresner ^10^, Adrienn Angyal ^93^, Luke R Green ^93^, Paul J Parsons ^93^, Rachel M Tucker ^93^, Rebecca Brown ^93^ and Max Whiteley ^93^.

**Software and analysis tools:**

James Bonfield ^99^, Christoph Puethe ^99^, Andrew Whitwham ^99^, Jennifier Liddle ^99^, Will Rowe ^41^, Igor Siveroni ^39^, Thanh Le-Viet ^70^ and Amy Gaskin ^69^.

**Visualisation:**

Rob Johnson ^39^.

**1** Barking, Havering and Redbridge University Hospitals NHS Trust, **2** Basingstoke Hospital, **3** Belfast Health & Social Care Trust, **4** Betsi Cadwaladr University Health Board, **5** Big Data Institute, Nuffield Department of Medicine, University of Oxford, **6** Brighton and Sussex University Hospitals NHS Trust, **7** Cambridge Stem Cell Institute, University of Cambridge, **8** Cambridge University Hospitals NHS Foundation Trust, **9** Cardiff and Vale University Health Board, **10** Cardiff University, **11** Centre for Clinical Infection & Diagnostics Research, St. Thomas’ Hospital and Kings College London, **12** Centre for Clinical Infection and Diagnostics Research, Department of Infectious Diseases, Guy’s and St Thomas’ NHS Foundation Trust, **13** Centre for Enzyme Innovation, University of Portsmouth (PORT), **14** Centre for Genomic Pathogen Surveillance, University of Oxford, **15** Clinical Microbiology Department, Queens Medical Centre, **16** Clinical Microbiology, University Hospitals of Leicester NHS Trust, **17** County Durham and Darlington NHS Foundation Trust, **18** Deep Seq, School of Life Sciences, Queens Medical Centre, University of Nottingham, **19** Department of Infection Biology, Faculty of Infectious & Tropical Diseases, London School of Hygiene & Tropical Medicine, **20** Department of Infectious Diseases, King’s College London, **21** Department of Microbiology, Kettering General Hospital, **22** Departments of Infectious Diseases and Microbiology, Cambridge University Hospitals NHS Foundation Trust; Cambridge, UK, **23** Department of Zoology, University of Oxford, **24** Division of Virology, Department of Pathology, University of Cambridge, **25** East Kent Hospitals University NHS Foundation Trust, **26** East Suffolk and North Essex NHS Foundation Trust, **27** Gateshead Health NHS Foundation Trust, **28** Genomics Innovation Unit, Guy’s and St. Thomas’ NHS Foundation Trust, **29** Gloucestershire Hospitals NHS Foundation Trust, **30** Great Ormond Street Hospital for Children NHS Foundation Trust, **31** Guy’s and St. Thomas’ BRC, **32** Guy’s and St. Thomas’ Hospitals, **33** Hampshire Hospitals NHS Foundation Trust, **34** Health Data Research UK Cambridge, **35** Health Services Laboratories, **36** Heartlands Hospital, Birmingham, **37** Hub for Biotechnology in the Built Environment, Northumbria University, **38** Imperial College Hospitals NHS Trust, **39** Imperial College London, **40** Institute of Biodiversity, Animal Health & Comparative Medicine, **41** Institute of Microbiology and Infection, University of Birmingham, **42** King’s College London, **43** Liverpool Clinical Laboratories, **44** Maidstone and Tunbridge Wells NHS Trust, **45** Manchester University NHS Foundation Trust, **46** Microbiology Department, Wye Valley NHS Trust, Hereford, **47** MRC Biostatistics Unit, University of Cambridge, **48** MRC-University of Glasgow Centre for Virus Research, **49** National Infection Service, PHE and Leeds Teaching Hospitals Trust, **50** Newcastle Hospitals NHS Foundation Trust, **51** Newcastle University, **52** NHS Greater Glasgow and Clyde, **53** NHS Lothian, **54** Norfolk and Norwich University Hospital, **55** Norfolk County Council, **56** North Cumbria Integrated Care NHS Foundation Trust, **57** North Tees and Hartlepool NHS Foundation Trust, **58** Northumbria University, **59** Oxford University Hospitals NHS Foundation Trust, **60** PathLinks, Northern Lincolnshire & Goole NHS Foundation Trust, **61** Portsmouth Hospitals University NHS Trust, **62** Princess Alexandra Hospital Microbiology Dept., **63** Public Health Agency, **64** Public Health England, **65** Public Health England,

Clinical Microbiology and Public Health Laboratory, Cambridge, UK, **66** Public Health England, Colindale, **67** Public Health England, Colindale, **68** Public Health Scotland, **69** Public Health Wales NHS Trust, **70** Quadram Institute Bioscience, **71** Queen Elizabeth Hospital, **72** Queen’s University Belfast, **73** Royal Devon and Exeter NHS Foundation Trust, **74** Royal Free NHS Trust, **75** Sandwell and West Birmingham NHS Trust, **76** School of Biological Sciences, University of Portsmouth (PORT), **77** School of Pharmacy and Biomedical Sciences, University of Portsmouth (PORT), **78** Sheffield Teaching Hospitals, **79** South Tees Hospitals NHS Foundation Trust, **80** Swansea University, **81** University Hospitals Southampton NHS Foundation Trust, **82** University College London, **83** University Hospital Southampton NHS Foundation Trust, **84** University Hospitals Coventry and Warwickshire, **85** University of Birmingham, **86** University of Birmingham Turnkey Laboratory, **87** University of Brighton, **88** University of Cambridge, **89** University of East Anglia, **90** University of Edinburgh, **91** University of Exeter, **92** University of Liverpool, **93** University of Sheffield, **94** University of Warwick, **95** University of Cambridge, **96** Viapath, Guy’s and St Thomas’ NHS Foundation Trust, and King’s College Hospital NHS Foundation Trust, **97** Virology, School of Life Sciences, Queens Medical Centre, University of Nottingham, **98** Wellcome Centre for Human Genetics, Nuffield Department of Medicine, University of Oxford, **99** Wellcome Sanger Institute, **100** West of Scotland Specialist Virology Centre, NHS Greater Glasgow and Clyde, **101** Department of Medicine, University of Cambridge, **102** Ministry of Health, Sri Lanka, **103** NIHR Health Protection Research Unit in HCAI and AMR, Imperial College London, **104** North West London Pathology, **105** NU-OMICS, Northumbria University, **106** University of Kent, **107** University of Oxford, **108** University of Southampton, **109** University of Southampton School of Health Sciences, **110** University of Southampton School of Medicine, **111** University of Surrey, **112** Warwick Medical School and Institute of Precision Diagnostics, Pathology, UHCW NHS Trust, **113** Wellcome Africa Health Research Institute Durban and **114** Wellcome Genome Campus.

## References

1 Public Health England. Investigation of novel SARS-COV-2 variant Variant of Concern 202012/01 Technical briefing 1. 2020.

2 Public Health England. Investigation of Novel SARS-COV-2 Variant Variant of Concern 202012/01 Technical Briefing 2. 2020.

3 The Lancet. Genomic sequencing in pandemics. Lancet 2021; 397: 445.

4 Davies NG, Barnard RC, Jarvis CI, et al. Estimated transmissibility and severity of novel SARS-CoV-2 Variant of Concern 202012/01 in England. medRxiv 2020. https://www.medrxiv.org/content/10.1101/2020.12.24.20248822v1.full-text.

5 Volz E, Mishra S, Chand M, et al. Transmission of SARS-CoV-2 Lineage B.1.1.7 in England: Insights from linking epidemiological and genetic data. DOI:10.1101/2020.12.30.20249034.

6 Peter Horby, Catherine Huntley, Nick Davies, John Edmunds, Neil Ferguson, Graham Medley, Andrew Hayward, Muge Cevik, Calum Semple. Update note on B.1.1.7 severity. NERVTAG, 2021.

7 Drew DA, Nguyen LH, Steves CJ, et al. Rapid implementation of mobile technology for real-time epidemiology of COVID-19. Science 2020; 368: 1362–7.

8 COVID-19 Genomics UK (COG-UK). An integrated national scale SARS-CoV-2 genomic surveillance network. Lancet Microbe 2020; 1: e99– 100.

9 Public Health England. Investigation of Novel SARS-COV-2 Variant Variant of Concern 202012/01 Technical Briefing 3. 2021.

10 NASA. NASA POWER Climate Data. https://power.larc.nasa.gov/ (accessed Jan 25, 2021).

11 Sudre CH, Murray B, Varsavsky T, et al. Attributes and predictors of Long-COVID: analysis of COVID cases and their symptoms collected by the Covid Symptoms Study App. DOI:10.1101/2020.10.19.20214494.

12 Varsavsky T, Graham MS, Canas LS, et al. Detecting COVID-19 infection hotspots in England using large-scale self-reported data from a mobile application: a prospective, observational study. The Lancet Public Health 2020; published online Dec 3. DOI:10.1016/S2468-2667(20)30269-3.

13 Public Health England. UK Covid-19 Dashboard. https://coronavirus.data.gov.uk/ (accessed Jan 19, 2021).

14 Walker AS, Sarah Walker A, Vihta K-D, et al. Increased infections, but not viral burden, with a new SARS-CoV-2 variant. DOI:10.1101/2021.01.13.21249721.

15 Office for National Statistics. Coronavirus (COVID-19) Infection Survey: characteristics of people testing positive for COVID-19 in England. 2021 https://www.ons.gov.uk/peoplepopulationandcommunity/healthandsocialcare/conditionsand diseases/articles/coronaviruscovid19infectionsinthecommunityinengland/characteristicsofpe opletestingpositiveforcovid19inengland27january2021.

16 Kissler SM, Fauver JR, Mack C, et al. Densely sampled viral trajectories suggest longer duration of acute infection with B.1.1.7 variant relative to non-B.1.1.7 SARS-CoV-2. DOI:10.1101/2021.02.16.21251535.

17 Hall V, Foulkes S, Charlett A, et al. Do antibody positive healthcare workers have lower SARS-CoV-2 infection rates than antibody negative healthcare workers? Large multi-centre prospective cohort study (the SIREN study), England: June to November 2020. bioRxiv. 2021; published online Jan 15. DOI:10.1101/2021.01.13.21249642.

18 Xie X, Zou J, Fontes-Garfias CR, et al. Neutralization of N501Y mutant SARS-CoV-2 by BNT162b2 vaccine-elicited sera. Cold Spring Harbor Laboratory. 2021; : 2021.01.07.425740.

19 Wu K, Werner AP, Moliva JI, et al. mRNA-1273 vaccine induces neutralizing antibodies against spike mutants from global SARS-CoV-2 variants. DOI:10.1101/2021.01.25.427948.

20 Muik A, Wallisch A-K, Sänger B, et al. Neutralization of SARS-CoV-2 lineage B.1.1.7 pseudovirus by BNT162b2 vaccine-elicited human sera. Science 2021; published online Jan 29. DOI:10.1126/science.abg6105.

21 Griffin J, Casey M, Collins Á, et al. Rapid review of available evidence on the serial interval and generation time of COVID-19. BMJ Open 2020; 10: e040263.

22 UK Government R estimates. https://www.gov.uk/guidance/the-r-number-in-the-uk (accessed Jan 21, 2020).

23 Coronavirus (COVID-19) Infection Survey, antibody data for the UK: 3 February 2021. Office for National Statistics, 2021.

24 Grubaugh ND, Hodcroft EB, Fauver JR, Phelan AL, Cevik M. Public health actions to control new SARS-CoV-2 variants. Cell 2021; published online Jan 29. DOI:10.1016/j.cell.2021.01.044.

25 Public Health England. Variants: distribution of cases data. 2021 https://www.gov.uk/government/publications/covid-19-variants-genomically-confirmed-case-numbers/variants-distribution-of-cases-data.

26 Baltrusaitis K, Santillana M, Crawley AW, Chunara R, Smolinski M, Brownstein JS. Determinants of Participants’ Follow-Up and Characterization of Representativeness in Flu Near You, A Participatory Disease Surveillance System. JMIR Public Health and Surveillance. 2017; 3: e18.

27 Griffith GJ, Morris TT, Tudball MJ, et al. Collider bias undermines our understanding of COVID-19 disease risk and severity. Nat Commun 2020; 11: 5749.

28 Zheng J, Zhou R, Chen F, et al. Incidence, clinical course and risk factor for recurrent PCR positivity in discharged COVID-19 patients in Guangzhou, China: A prospective cohort study. PLOS Neglected Tropical Diseases. 2020; 14: e0008648.

29 Kifer D, Bugada D, Villar-Garcia J, et al. Effects of environmental factors on severity and mortality of COVID-19. MedRxiv 2020. https://www.medrxiv.org/content/10.1101/2020.07.11.20147157v3.full-text.

